# DAT-SPECT profiling for biological definition of two prospective Parkinson’s disease cohorts

**DOI:** 10.1101/2025.03.11.25323734

**Authors:** Rachele Malito, Chiara Meneghini, Alice Galli, Luca Gallo, Pierfrancesco Mitrotti, Paola Dimartino, Marianna Inglese, Cinzia Zatti, Alessandro Lupini, Andrea Pilotto, Alessandro Padovani, Micol Avenali, Enza Maria Valente, Cristina Tassorelli, Silvia Paola Caminiti

## Abstract

**Background:** There is a need for simplified classification systems capable of effectively stratifying Parkinson’s disease (PD) in routine clinical practice.

**Objectives:** Identification of PD subtypes integrating neuroimaging and clinical features, widely used to support clinical diagnosis.

**Methods:** We included, from Parkinson’s Progression Markers Initiative (PPMI), 249 *de novo* early diagnosed PD patients. Two step clustering analysis was run on ^123^I-FP-CIT-SPECT striatal uptake [D+/D] and motor impairment [M+/M]. The emerging subgroups were evaluated for demographic, clinical, cerebrospinal fluid biomarkers, brain morphometry and longitudinal clinical progression. Mediation analysis evaluated the effect of biomarkers on the relationship between subgroups and cognitive decline. We validated the proposed classification algorithm through an independent validation cohort (n=84).

**Results:** Four distinct subtypes emerged: [D+/M+]: poorer memory performance, greater Aβ_1-42_ and α-synuclein pathology and atrophy in the inferior temporo-occipital cortex. At follow-up D+/M+ showed faster progression of motor disability, motor complications and cognitive decline. [D/M]: higher levels of anxiety and gray matter volume reductions in the precuneus, fusiform gyrus and the precentral gyrus. [D/M+]: marked by pronounced rigidity and apathy, atrophy in motor and sensorimotor areas, and a more rapid progression of rigidity. [D+/M]: severe impulsiveness, lower working memory performance and thalamic atrophy.

Aβ_1-42_ showed significant mediational effect on the relationship between D+/M+ and cognitive decline.

The validation cohort supported the clinical findings observed in the PPMI cohort.

**Conclusions:** PD can be categorized into distinct subgroups using information widely available in clinical and research settings. These data offer valuable insights into the underlying co-pathology, disease progression and severity.

**Graphical abstract:** 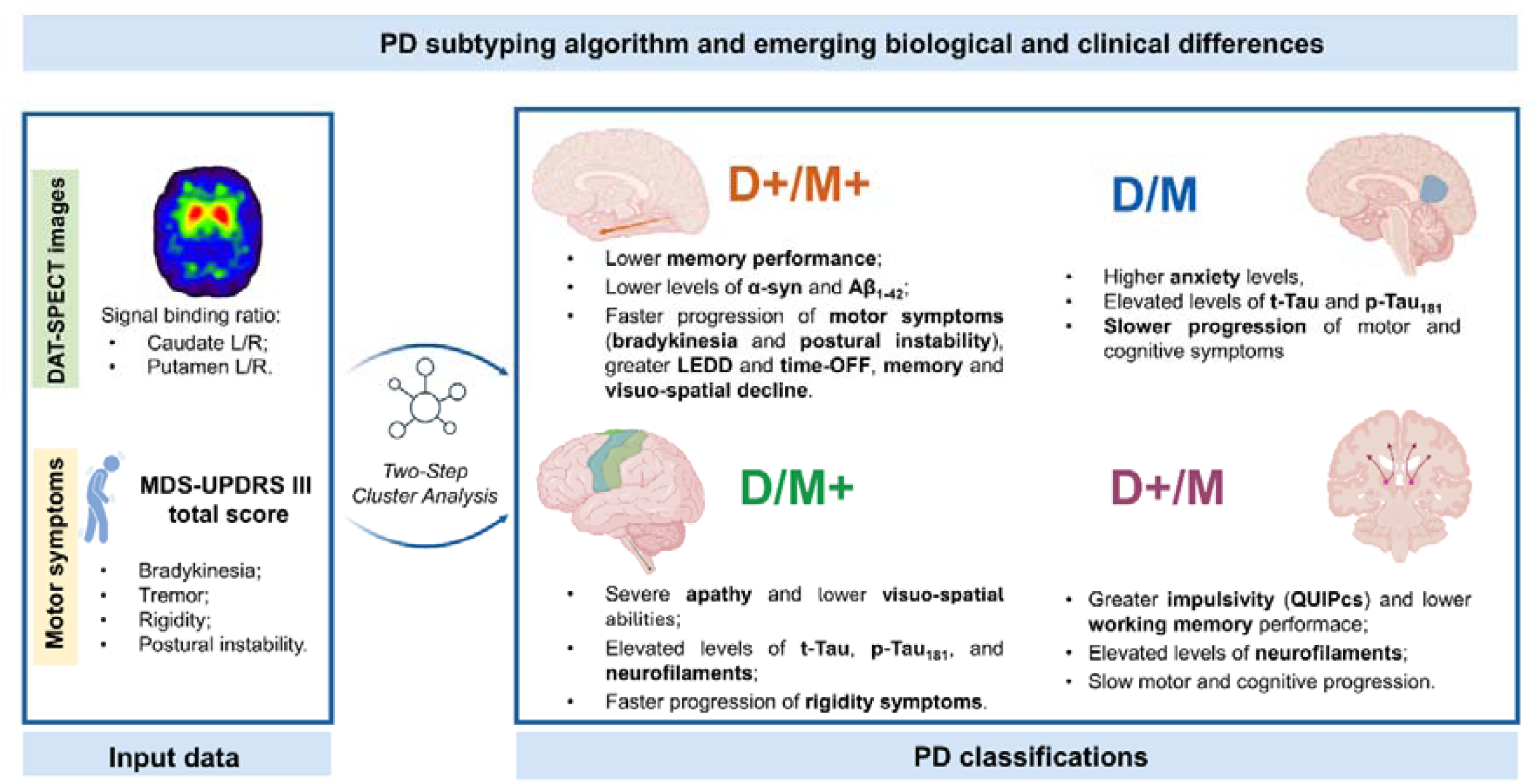

## 1. Introduction

Parkinson’s disease (PD) is currently defined by the presence of cardinal motor features, specifically bradykinesia along with either resting tremor or rigidity, or both.^1^ The movement disorder society (MDS) highlights the importance of a nigrostriatal dopaminergic deficit to make a diagnosis of PD.^1^ Dopamine transporter single-photon emission computed tomography (*DAT-SPECT*) is the main neuroimaging technique used in both clinical research and practice to assess presence and the degree of nigrostriatal dopaminergic dysfunction.^2^ Recently, together with dopaminergic denervation, the assessment of alpha-synuclein (α-syn), through seeding amplification assay (SAA) techniques, has allowed the development of new biological research criteria for synucleinopathy.^3,4^ The technical progress allowed for the *in vivo* detection of misfolded α-syn, hence offering the possibility of redefining PD according to both biological and clinical features.^4^

However, PD clinical presentation and prognosis are highly heterogeneous,^5,6^ and PD is frequently accompanied by a diversified pool of concomitant pathologies as well as neuropsychiatric manifestations^7^, which are probably linked to the degeneration of other structures and systems aside from the nigrostriatal system. Taken together, these findings challenge the view of PD as a unitarian disorder; thus, stimulating the interest towards the identification of PD subtypes.^8^

Several studies employed hypothesis-free data-driven approaches to identify PD subtypes based on demographic and clinical information.^6,9^ However, the majority of studies either considered heterogeneous cohorts of patients^9^ or included small sample of *de novo* PD patients or considered a great number of variables, some of which not easily available in clinical routine.^10^ For these reasons, the proposed stratifications of PD patients, leaves open the possibility that these subtypes do not truly represent real distinct disease subtypes. Moreover, previous studies failed to highlight the importance of the *DAT-SPECT* uptake reduction, a well-established predictor of PD phenotypic presentation.^1,11^ Indeed, various studies suggest that the pattern and severity of dopaminergic damage has a strong prognostic and predictive value of disease progression.^12,13^

Here, we aim to explore the relationship between dopaminergic denervation and motor impairment severity, focusing on cases with mismatched DAT-SPECT and motor presentations.

## 2. Materials and methods

### 2.1 Study participants

Data used in this study was acquired from the Parkinson’s Progression Markers Initiative (PPMI) database. Data acquired between July 1, 2010, and May 1, 2016 were downloaded from the PPMI public database (www.ppmi-info.org/access-data-specimens/downloaDdata), RRID:SCR_006431. For up-to-date information on the study, visit www.ppmi-info.org.^14^

Our sample consisted of 249 drug-naïve PD patients (mean age±SD: 62.95±9.04 years; sex [F/M]: 83/166) with a disease duration of ≤ 2 years and (**see Supplementary Materials**). Patients underwent an average of 8 years of clinical follow-up (mean±SD: 7.71; range: 1–13 years), during which comprehensive demographic and clinical assessments were conducted (**Supplementary Materials**).

### 2.2 *DAT-SPECT* imaging analysis

All participants underwent *DAT-SPECT*; reconstructed images are available on the PPMI site (www.ppmi-info.org/data). Images pre-processing was conducted with Statistical Parametric Mapping (SPM12). Each reconstructed image passed a quality check before being included. After images spatial normalization(http://www.nitrc.org/projects/spmtemplates)^15^ specific binding ratio (SBR) was obtained considering lateral superior occipital cortex as reference. ^16^

ROIs were selected from the Automated Anatomical Labelling (AAL) atlas.^17^ The posterior fornix was considered the boundary between anterior and posterior putamen. SBR in the posterior putamen were used to calculate asymmetry index (AI) through Walker et al formula. ^18^

### 2.3 CSF biomarkers

Baseline data regarding seeding aggregation activity (SAA) and Cerebrospinal fluid (CSF) biomarkers for each patient were acquired from the PPMI biospecimen database to highlight differences among clusters and subgroups. For further information see **Supplementary Materials**.

### 2.4 Gray matter volume analysis

#### Sample characteristics

We selected T1-weighted Magnetic resonance imaging (MRI) from the PPMI database. Further details about T1-MRI scans acquisitions are available: https://www.ppmi-info.org/sites/default/files/docs/archives/PPMI2.0_MRI_TOM_Final_FullyExecuted_v2.0_20200807.pdf. High resolution T1-weighted MRI were available for 219 PD patients and 128 healthy control (HC) subjects (mean age±SD: 58.91±11.6 years; sex [M/F]: 93/47).

The HC group had no first-degree family history of PD, no history of neurological disorders, stable Movement Disorders Society-Unified Parkinson’s Disease Rating Scale-Part III (MDS-UPDRS part III) scores, no PD diagnosis over 6-year follow-up and were negative for the main PD related mutations. HC were matched to each subgroup for age, sex and numerosity [HC matched D+/M+ n=35; HC matched D/M n=75; HC matched D/M+ n=35; HC matched D+/M n=67].

#### Voxel-based morphometry (VBM)

All MRI images were processed using CAT12 (https://neuro-jena.github.io/cat). The standard segmentation procedure was employed to differentiate gray matter, white matter and CSF. The result of VBM analysis were normalized and modulated gray matter segmentations. After quality-control check (subjects with processing quality ≤ B-were excluded; n PD=6, n HC=4), total intracranial volume (TIV) values were extracted. Images were smoothed with a Gaussian kernel of 6 mm.

#### MRI between group comparison

A two-sample t-test was conducted in CAT12 to compare D+ and D clusters and each subgroup with an age and sex-matched HC subgroup. TIV, age at MRI, sex and years of education were included as covariates. Voxel-wise significance threshold was set at two-tailed p < 0.005 with a minimum cluster extent of 100 voxels. JuBrain Anatomy Toolbox (https://github.com/inm7/jubrain-anatomy-toolbox) allowed anatomical localization.

### 2.5 Statistical analysis

We ran a two-step cluster analysis twice to explore empirical clusters within the PD cohort considering, first, *DAT-SPECT* SBR of left and right caudate and putamen. Then, clustering was performed considering MDS-UPDRS III total score and motor sub-scores: bradykinesia, postural, rigidity and tremor (Motor feature, “M”). Next, we combined the two classifications (**Supplementary** Fig. 1).

Chi-squared, one-way ANOVA and Kruskal-Wallis H tests were performed to compare demographic features of clusters and subgroups. Moreover, patients were classified based on a previously proposed classification^10^ into “mild motor-predominant”, “intermediate” and “diffuse malignant”.

Clinical, CSF and *DAT-SPECT* characteristics (dependent variables) were evaluated using linear and logistic regression models considering clusters and subgroups as independent variables. Analyses were performed adjusting for sex, age and disease duration at baseline. Multiple comparisons were adjusted with the Benjamini-Hochberg test.

The longitudinal rate of change for D+/D in a) **motor**, i.e. Levodopa equivalent daily dose (LEDD), MDS-UPDRS part III, bradykinesia, tremor, rigidity, and postural sub-scores; MDS-UPDRS part IV sub-scores: total hours with dyskinesia, total OFF hours, functional impact of fluctuations, complexity of motor fluctuations, and b) **cognitive performance**, i.e., Montreal Cognitive Assessment (MoCA) score, Benton Judgment of Line Orientation (BJLO), Hopkins Verbal Learning Test (HVLT), Letter Number Sequencing (LNS), Symbol Digit Modality (SDM) and semantic fluency, was compared through a mixed linear model, adjusting for age, sex and education. The same analysis was performed to investigate the rate of disease progression across the four subgroups. In this case mixed linear model was calculated comparing each single subgroup against the whole sample and by performing pair-to-pair comparisons between subgroups. Mixed linear models is able to operate with missing values, correlated data, different numbers of observations and inconsistent observations intervals.

We conducted a mediation analysis ^19^ to investigate whether the relationship between D+/M+ and longitudinal changes in MoCA slope were mediated by CSF biomarkers. Individual MoCA slope were obtained through linear regression model, considering MoCA scores as the dependent variable and time as the independent variable, adjusting for age, sex and education. All pathways were tested using linear regression modelling, reported as adjusted β, 95% CI, and p-value (**Supplementary Table 1)**. Two-tailed p-values were considered for all statistical analyses. All statistical analyses were performed using R software, version 3.6.3.^23,24^

### 2.6 Sensitivity analysis

We used internal validation data derived from a research protocol approved by the Ethics Committee of Brescia Hospital, Brescia, Italy (DNA study, NP 1471) to validate the finding obtained with the PPMI cohort (**see Supplementary Materials, Methods**).

## 3. Results

### 3.1 Cluster analysis

The *DAT-SPECT* classification identified two clusters: D+ (severe dopaminergic denervation) and D (mild dopaminergic damage) with a sample size of 120 (48.2%) and 129 (51.8%), respectively (Silhouette coefficient equal to 0.6).

The motor classification provided two clusters: M+, with relatively higher scores in MDS-UPDRS part III total score and subitems, and M cluster, displaying milder motor deficit. Clusters have a sample size of 88 (35.3%) and 161 (64.7%), respectively (Silhouette coefficient = 0.5). As expected, M+ displayed more severe motor impairment in terms of MDS-UPDRS part III total score compared to M (**Supplementary Table 2**).

By combining the severity of motor symptoms and the dopaminergic damage 4 subgroups emerged: n=44 (17.7%) **D+/M+**; n=44 (17.7%) **D/M+**; n=76 (30.5%) **D+/M**; n=85 (34.1%) **D/M**. Note that all subgroups of PD patients exhibited significantly lower uptake in the caudate and putamen and MDS-UPDRS part III total score compared to HC group (**Supplementary** Fig. 2).

### 3.2 Differences in baseline and follow-up data

D+/D clusters were similar for age, age at onset, sex, education and disease duration (**Supplementary Table 3**). At baseline, the D+ cluster exhibited severe postural instability and lower memory performances (**Fig. 1A** and **Supplementary Table 4**). This cluster also showed greater α-syn and p-Tau_181_/α-syn ratio pathology (**Fig. 1A** and **Supplementary Table 5**). When directly compared to the D cluster, D+ exhibited GMV reductions in bilateral thalamus, left caudate nucleus, superior temporal gyrus, temporal pole, precentral gyrus, middle frontal gyrus (orbital part), right inferior frontal gyrus (triangular part).

**Fig. 1.**
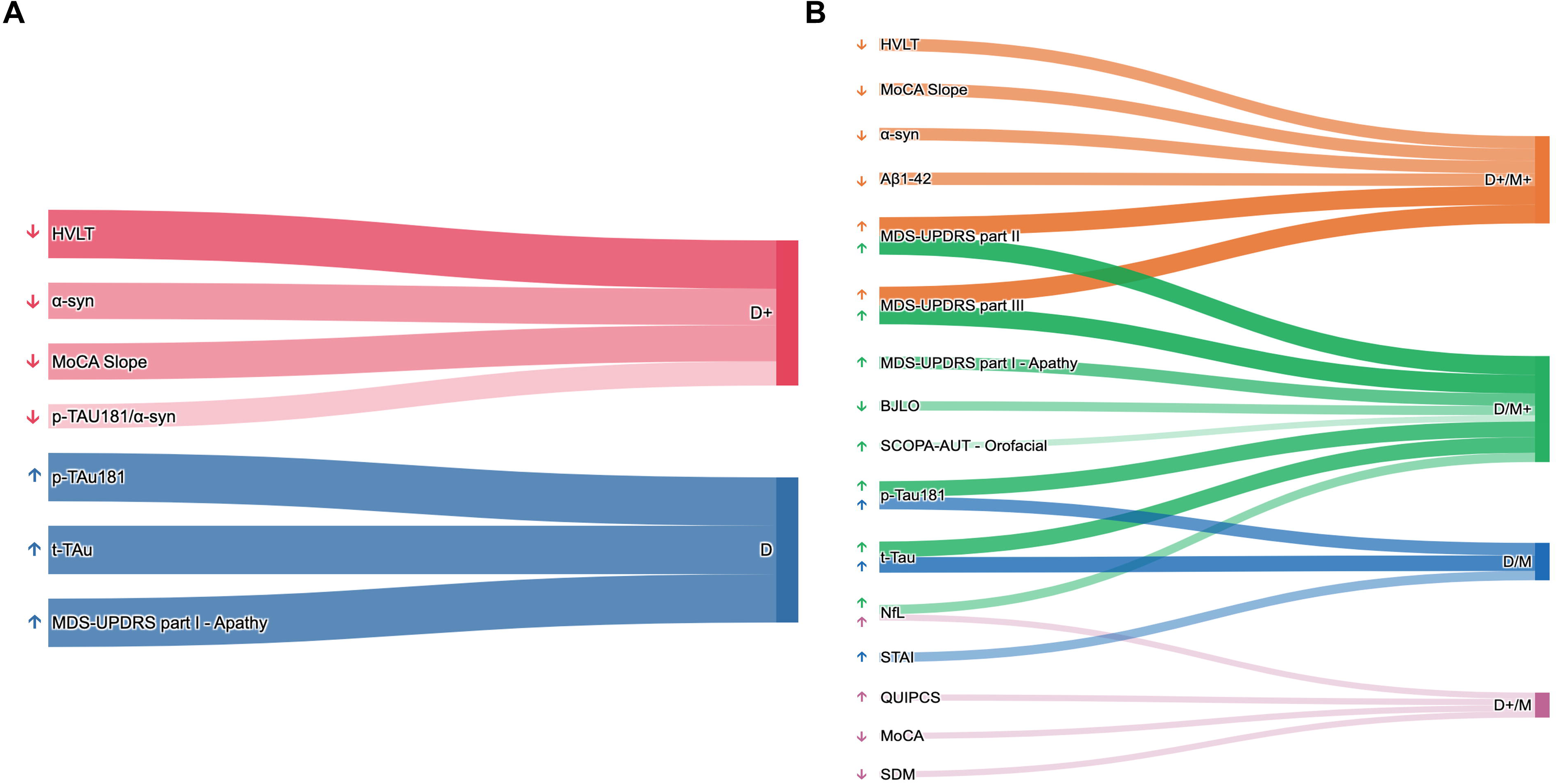
Sankey Diagram representing associations between clinical, neuropsychological, and biological variables across Parkinson’s Disease clusters and subgroups. Representation of the associations between clinical, neuropsychological and biological variables and (**A**) D+/D and (**B**) the four Parkinson’s disease subgroups. Line thickness and opacity correlates with the p-values of the associations, with thicker and darker lines indicating stronger significance. <u>Abbreviations</u>: α-synuclein (α-syn); Aβ_1-42_ (Aβ_1-42_); Montreal Cognitive Assessment (MoCA); Hopkins Verbal Learning Test (HVLT); Movement Disorders Society - Unified Parkinson’s Disease Rating Scale (MDS-UPDRS); Benton Judgment of Line Orientation (BJLO); Scales for Outcomes in PD Autonomic (SCOPA-AUT); Phosphorylated Tau (p-Tau_181_); Total Tau (t-Tau); Neurofilament light Chain (NfL); State Trait Anxiety Inventory (STAI).

At follow-up, the D+ cluster displayed faster progression of bradykinesia (1.5 points) and more severe postural instability (0.5 points per year). Additionally, the D+ cluster resulted more prone to develop motor complication in terms of longer time spent in the OFF state (0.73 point per year). Cognitively, the D+ cluster exhibited a faster decline on HVLT memory performance (−2.4 points per year) (**Supplementary Table 6**). The more severe cognitive decline of the D+ cluster was confirmed by the steeper MoCA slope compared to the D cluster (**Supplementary Table 4**).

Instead, the D cluster exhibited greater MDS-UPDRS part I apathy sub-score along with greater total and p-Tau_181_ CSF levels (**Fig. 1A** and **Supplementary Table 4-5**). Compared to the D+ cluster, the D cluster didn’t show evidence of atrophy. At follow-up, the D cluster exhibited a faster progression of tremor, (0.8 points). See **Supplementary Table 6**. The four subgroups were similar for age, age at onset, sex, education and disease duration (**Supplementary Table 7**).

At baseline, the D+/M+ subgroup was characterized by lower memory performance, together with greater bradykinesia, postural instability and MDS-UPDRS part II compared to M subgroups (**Figure 2B** and **Table 1**). Moreover, the D+/M+ subgroup showed more pathological levels of α-syn and Aβ_1-42_ compared to D subgroups (**Fig. 1B** and **Supplementary Table 8**). Additionally, the D+/M+ subgroup, compared to HC, exhibited GMV loss in bilateral fusiform gyrus and left lingual gyrus (**Fig. 2**).

**Fig. 2.**
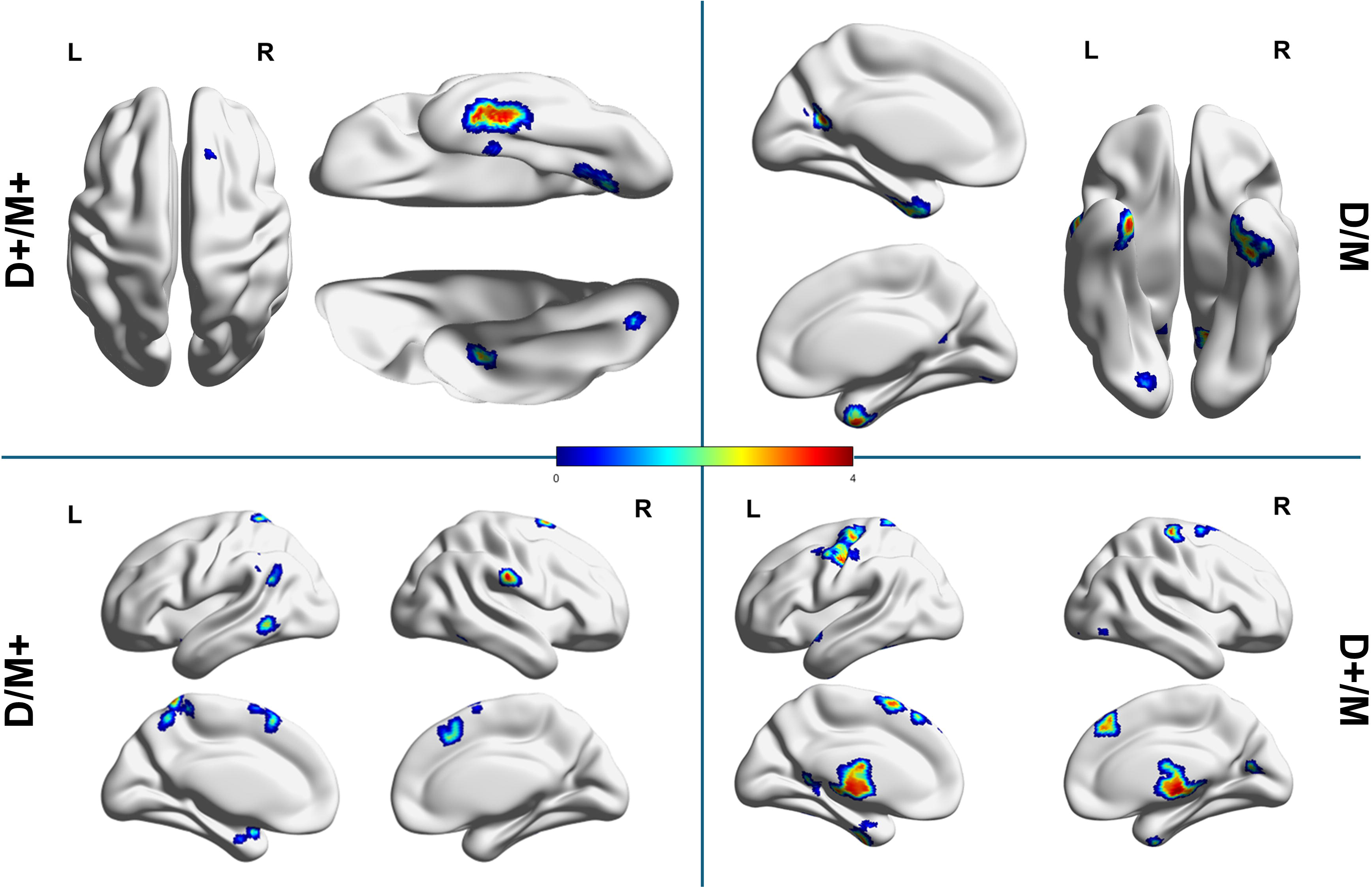
VBM analysis of Gray Matter Abnormalities in Parkinson’s Disease Subgroups Using MRI. Segmented T1-weighted images of the four subgroups were compared to HC (matched for numerosity, age at MRI acquisition, and sex), with adjustments for total intracranial volume (TIV), age at MRI acquisition, sex, and years of education. p-value = 0.005 (uncorrected) and extent threshold k = 100. BrainNet Viewer (http://www.nitrc.org/projects/bnv/) was used for rendering. ^50^

**Table 1.**
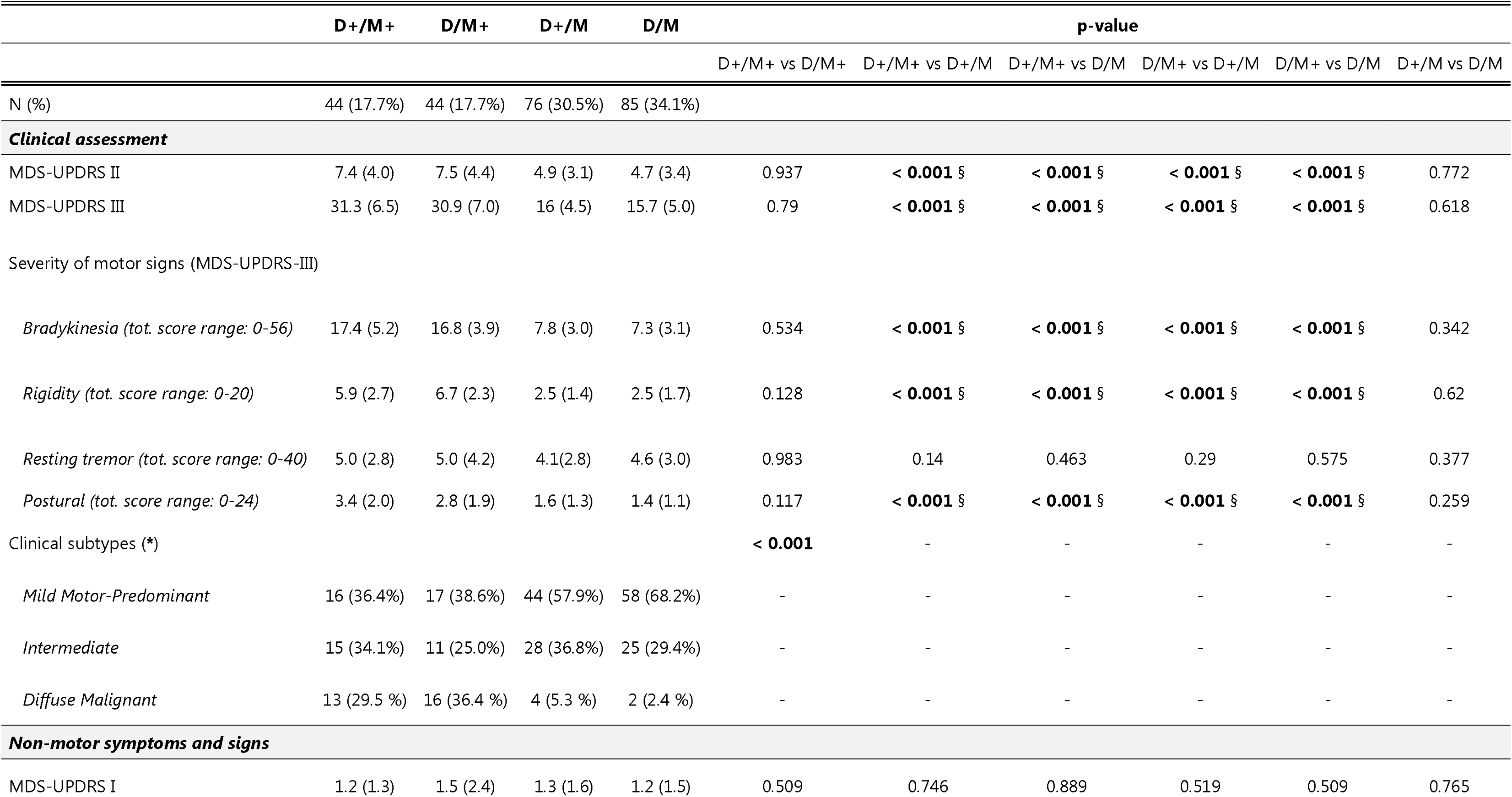
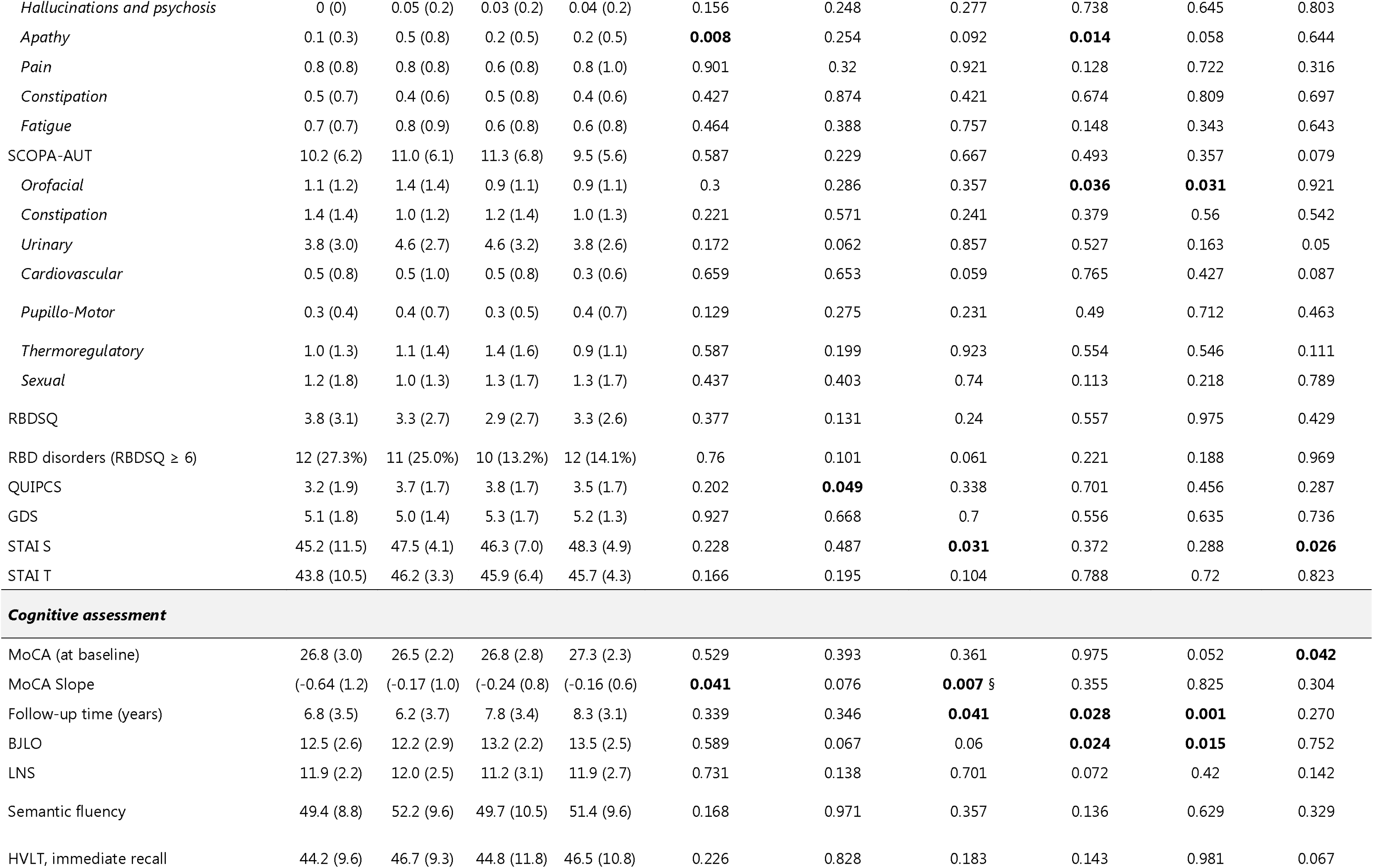
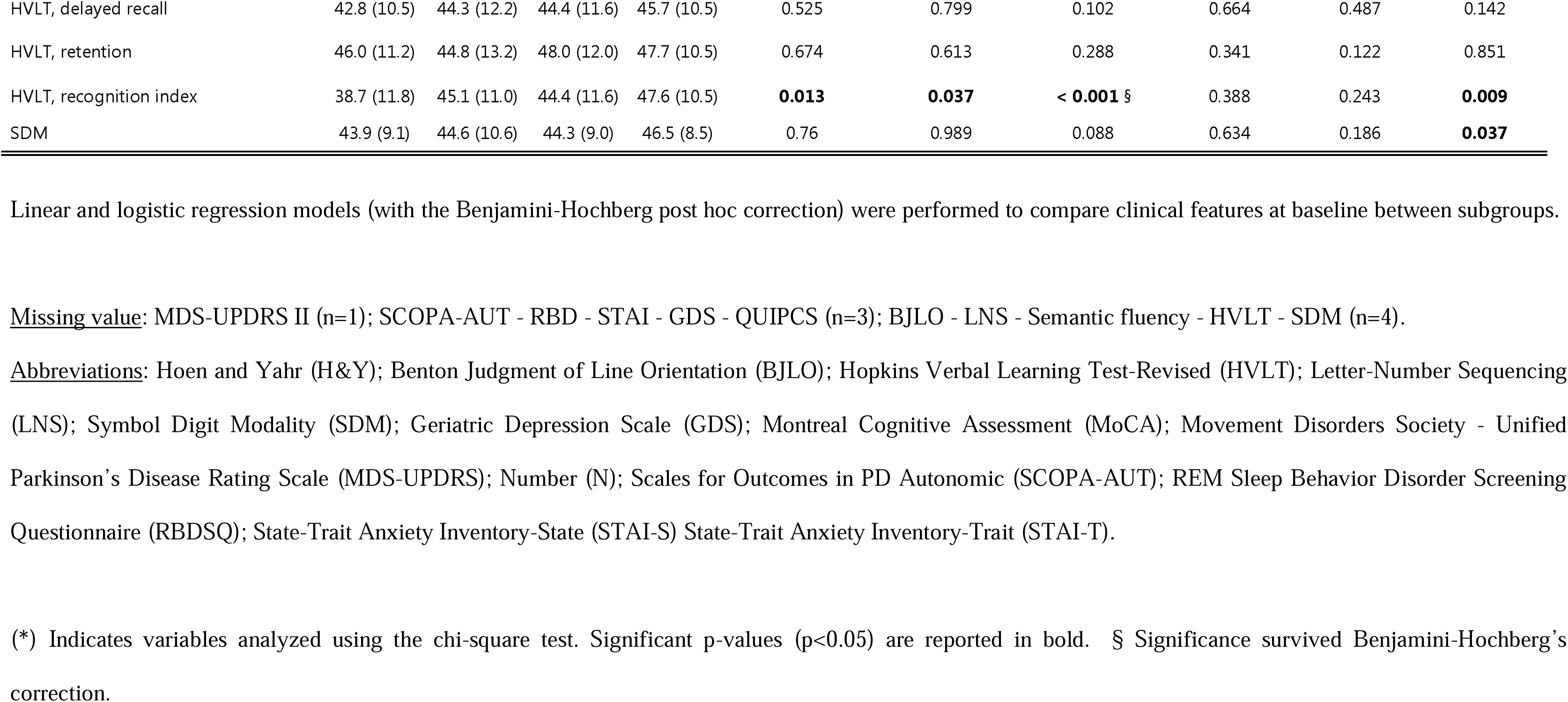
Comparison of the baseline clinical features among the four subgroups of Parkinson’s disease population.

At follow-up, the D+/M+ subgroup showed a marked increase in longitudinal LEDD values compared to the other subgroups (62.6 points per year) and greater annual rate of change in MDS-UPDRS part III motor scores (8.6 points per year). Notably, we observed more severe bradykinetic and postural symptoms, with an annual increase rate of 6.23 and 1.33 points, respectively, as well as longer duration of dyskinesia (1.1 points per year) and extended time spent in the OFF state (0.9 points per year). Cognitively, the D+/M+ group exhibited a faster annual decline in MoCA scores (−1.02 points per year). This decline was particularly pronounced for visuospatial abilities (−1.24 points per year), and memory (−3.17 points per year). See **Supplementary Table 9-10** for comparison across subgroups and **Fig. 3**.

**Fig. 3.**
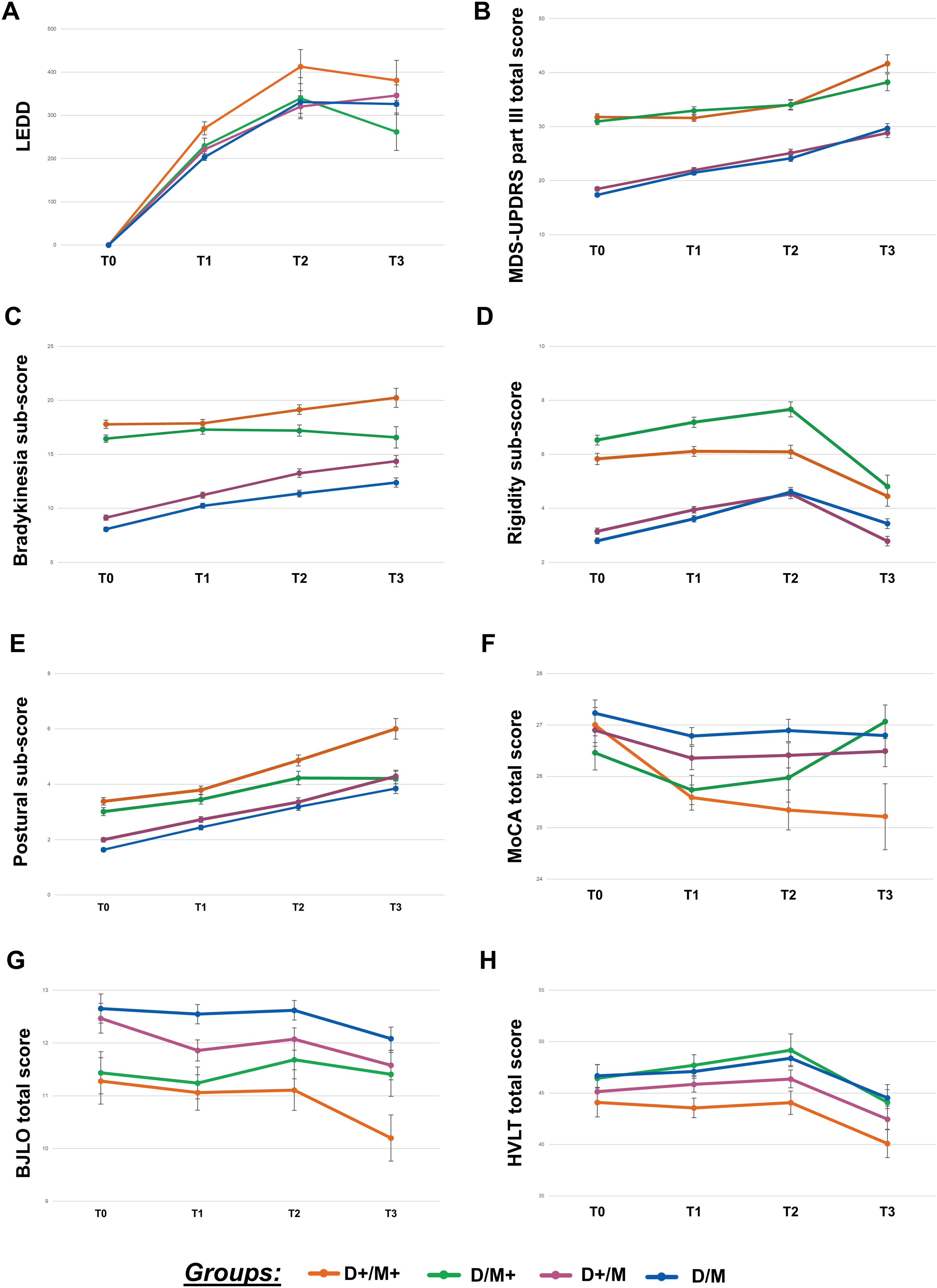
Temporal Trends in Cognitive Scores Across Four Parkinson’s Disease Subgroups. Lines depicting changes across different time points: T0 = baseline, T1 = 1-3 years of follow-up, T2 = 4-6 years of follow-up, and T3 = 7-13 years of follow-up. The analysis was performed using a mixed linear model. <u>Abbreviations</u>: Levodopa equivalent daily dose (LEDD); Movement Disorder Society - Unified Parkinson’s Disease Rating Scale (MDS-UPDRS); Montreal Cognitive Assessment (MoCA); Benton Judgment of Line Orientation (BJLO); Hopkins Verbal Learning Test (HVLT).

The D/M subgroup revealed the most benign presentation of the disease (**Fig. 1B** and **Supplementary Table 7**). Consistently, 68.2% of subjects in the D/M subgroups were classified as “mild motor-predominant phenotype” (**Table 1**). However, the D/M subgroup displayed higher state anxiety score and higher t-Tau and p-Tau_181_ levels compared to D+ subgroups (**Fig. 1B**, **Table 1 and Supplementary Table 8**). The D/M subgroup showed brain atrophy in precuneus, anterior fusiform gyrus and right precentral gyrus compared to HC (**Fig. 2**).

Of note, we found a subgroup showing prominent motor impairment despite a mild dopaminergic deficit, i.e. D/M+. D/M+, at baseline, showed more severe rigid symptomatology, lower visuospatial ability scores, higher MDS-UPDRS part II scores and higher scores at the Scales for

Outcomes in PD Autonomic (SCOPA-AUT) - Orofacial sub-score - compared to M subgroups as well as greater MDS-UPDRS part I - Apathy compared to D+ subgroups (**Fig. 1B** and **Supplementary table 7**). D/M+ subgroup showed higher levels of t-Tau, p-Tau_181_ and NfL (**Fig. 1B** and **Supplementary Table 8**). Finally, the D/M+ subgroup, compared to HC, showed GMV reduction involving parietal regions (left superior parietal gyrus, supramarginal gyrus, angular gyrus and right postcentral gyrus), supplementary motor area, right inferior temporal gyrus, left middle temporal gyrus and temporal pole (**Fig. 2**).

At follow-up, the D/M+ subgroup exhibited more severe rigidity symptoms (+2.55 points per year) compared to the rest of the cohort See **Supplementary Table 9-10** and **Fig. 3**.

On the other hand, the subgroup characterized by more prevalent dopaminergic than motor deficit, i.e. D+/M, showed slightly greater score in the Questionnaire for Impulsive-Compulsive Disorders in Parkinson’s Disease (QUIPCS) compared to D+/M+ subgroup, as well as slightly lower performances in MoCA and Symbol Digit Modalities test (SDM) compared to the D/M subgroup (**Fig. 1B** and **Table 1**). Additionally, D+/M showed greater NfL values compared to the D/M subgroup (**Fig. 1B and Supplementary table 8**). Finally, the D+/M subgroup showed significant loss of GMV in bilateral thalamus, right precentral gyrus, left postcentral gyrus, temporal pole and superior frontal gyrus compared to HC (**Fig. 2**).

### 3.3 Mediation analysis

The mediation analysis met all steps. Step 1: Only the D+/M+ subgroup was linked to MoCA slope decline. Steps 2-3: Several biomarkers correlated with decline, but only Aβ1-42 predicted D+/M+. Step 4: Aβ1-42 showed a moderate mediational effect (PM: 13%) (**Supplementary** Fig. 3 and **Supplementary Table 11**).

### 3.4 Sensitivity Analysis

The validation cohort confirmed the findings obtained with the PPMI cohort. For more details see **Supplementary Materials, Methods**.

## 4. Discussion

Here, we utilized *DAT-SPECT* imaging, a widely available clinical routine biomarker for supporting PD diagnosis, to systematically identify two distinct clusters (D/D+), reflecting milder and more severe dopaminergic damage, respectively. Importantly, the imaging data were acquired at baseline, in an early clinical phase, and in the absence of treatment, ensuring an unbiased assessment of dopaminergic denervation. Furthermore, we integrated the severity of dopaminergic denervation with motor impairment to define specific PD subgroups.

The groups identified at baseline, despite having similar disease duration, age at disease onset and SAA positivity, showed different clinical and biomarkers features and rates of PD progression. The recent development of SAAs techniques allowed for the *in vivo* detection and measurement of misfolded α-syn.^4,20^ However, as also suggested by the current findings, in PD, SAAs positivity is not informative for disease phenotypic stratification. The presence of co-pathology, the degree of dopaminergic denervation and the contribution of non-dopaminergic systems, significantly contribute to the phenotypic manifestation of the disease.^7^

Of note, the here proposed classification of PD patients, which aims at highlighting the essential role of dopaminergic damage in determining the phenotypic manifestation of the disease, does not overlap with previously proposed clinical stratifications. Indeed, when categorizing our PD patients into one of the three clusters previously proposed ^10^, we observed only partial concordance with our clustering stratification. Notably, our classification demonstrated a stronger alignment with motor symptom severity (M+/M-), potentially due to the extensive number of clinical variables incorporated in the latter algorithm. This broader variable selection may have facilitated the identification of PD subtypes reflecting distinct disease phenotypes, albeit with limited representation of the underlying neuropathological processes.

By focusing on dopaminergic denervation, our analysis highlighted that the D+ cluster was associated with more severe postural instability and greater memory impairment. This is coherent with the increased dopaminergic damage observed in the postural instability gait difficulty subtype and the significant association between postural instability and cognitive decline.^21,22^ At follow up, the D+ cluster exhibited a faster progression of bradykinesia, tremor and cognitive impairment along with higher probability of developing motor complication. Additionally, the comparison of MRI scan between D+ and D revealed significant GMV reductions in the thalamus and fronto-temporal regions. ^23,24^

On the other hand, the D cluster was mainly characterized, at baseline, by the presence of apathy, no evidence of brain atrophy, and a faster progression of tremor, at follow-up. These last features, in particular the presence of tremor and reduced atrophy, are in favor of a benign course of the disease.^25,26^ Hence, the hereby proposed stratification of PD patients, based only on dopaminergic features, is able to discriminate between a severe and benign PD phenotype.

By integrating classifications derived from clinical and instrumental assessments, we stratified four subgroups characterized by unique clinical phenotypes, biomarker expression profiles, and trajectories of cognitive and motor progression over time. Notably, the D+/M+ subgroup exhibited the most severe disease phenotype, marked by pronounced motor dysfunction—particularly bradykinesia and postural instability—alongside significant memory deficits and dopaminergic degeneration. The severity of this subgroup was also confirmed in an independent cohort of patients. These observations strongly confirmed previous evidence proposing both severe motor dysfunction and dopaminergic denervation at baseline as the cause of a more malignant PD phenotype.^10^ Furthermore, the presence of memory deficits and the focal reduction in GMV observed in the inferior occipito-temporal cortex of this subgroup are consistent with prior studies linking atrophy in these regions to development of dementia.^27,28^ These morphological changes have been linked to posterior cholinergic degeneration in PD, with increased degeneration observed in case of dementia, consistent with the proposed “dual-syndrome hypothesis”.^29,30^ We found that Aβ_1-42_ has a mediational effect on the association between the D+/M+ subgroup and cognitive decline. Consistent with our observations, the literature has extensively reported the predictive value of cerebrospinal fluid Aβ_1-42_ for global cognitive decline, domain-specific cognitive decline, motor function and autonomic function in patients with PD.^31^

At follow-up, the severity of initial motor symptoms and dopaminergic loss not only predicts faster progression of motor impairment but also correlates with an increased dependency on dopaminergic therapies over time. This subgroup is therefore expected to require more intensive healthcare resources. Furthermore, the D+/M+ subgroup, along with the rapid motor worsening of bradykinesia and postural instability, showed a deterioration in memory and visuospatial skills. These findings confirm the relationship between axial symptoms and increased visuospatial impairment, as well as the likely involvement of the lingual and fusiform regions visual cortical components with visual perceptual dysfunction.^32,33^

Conversely, the D/M subgroup revealed only a mild clinical picture, mainly characterized by the presence of anxiety. Even at follow-up, the D/M subgroup demonstrated the most benign prognosis. The comparison of MRI scans of this subgroup with controls revealed a sizable GMV loss in the precuneus, a structure linked to anxiety in PD.^34^

The higher p-Tau_181_ and t-Tau CSF levels we observed in the D/M subgroup at baseline, confirmed previous evidence suggesting the existence of a negative correlation between the levels of dopaminergic denervation and CSF Tau proteins.^35^ Further study investigating tauopathy, and the combined effect of α-syn and tauopathy on PD dopamine and clinical manifestations are needed. Despite a less severe dopaminergic denervation, the D/M+ subgroup exhibited a relatively more compromised motor profile, specifically characterized by heightened rigidity.

The D/M+ group showed widespread GMV loss in motor and sensorimotor regions. SMA is involved in motor sequencing, planning, and learning.^36,37^ Supramarginal/Postcentral gyrus adapt motor actions. ^38^ Angular gyrus monitors actions and discrepancies.^39,40^ Superior parietal gyrus aids motor planning and visuomotor coordination. ^41,42^

Moreover, rigidity, that characterize the D/M+ subgroup, has recently been studied through biomechanical and neurophysiological measures, suggesting a partial overlap between the neural circuits involved in long-latency reflexes and the pathways contributing to “objective rigidity” in PD.^43^ It has been proposed that the spino-cerebello-reticulo-spinal pathway may drive parkinsonian rigidity, potentially explaining the observed severity of motor impairment even in cases of relatively mild striatal dopaminergic damage.^43^

Finally, the D+/M subgroup was characterized by the presence of non-motor symptoms, such as Impulse Control Disorders (ICDs), cognitive and executive deficits. ICDs occur in approximately 19.1% of patients with early-stage PD, often persisting or worsening over time, particularly with dopaminergic medication use.^44^ PDICD cases appear to be associated with more severe involvement of the frontal, mesolimbic, and motor circuits.^45^ However, the lack of prospective studies limits our understanding of the progression and prognosis of PDICD.

MRI comparisons between this subgroup and HC revealed severe thalamic atrophy. The thalamus plays a crucial role in cortico-basal ganglia-cortical circuits.^46^ The thalamic abnormalities observed in this subgroup may be explained by dopamine depletion in the basal ganglia, leading to increased inhibitory drive from the globus pallidus/substantia nigra reticulata to thalamic neuronal activity.^46,47^ A recent study by Parker at al.,^48^ using optogenetics and recordings from striatal medium spiny neurons in transgenic mouse brain slices, demonstrated that dopamine depletion disrupts basal ganglia-thalamic communication, reducing spontaneous motor activity in parkinsonian mice. However, this dopamine-centered perspective overlooks the important role of noradrenergic inputs from the locus coeruleus, which strongly influence thalamic neuronal activity.^48,49^ Pifl et al.,^46^ in their study on post-mortem human brain tissue, found a severe reduction of noradrenaline in most thalamic nuclei in PD, comparable to the dopamine loss observed in the same patients in the caudate and putamen. Since noradrenaline deficiency impairs both the efficiency and accuracy of thalamic information processing, its depletion is expected to significantly impair signal transmission in the PD thalamus.^46^

## 5. Conclusion

By means of a hypothesis-free data driven approach, we stratified two independent cohorts of *de novo* PD patients into four different subtypes. The identified subgroups displayed differences in clinical manifestations, biological features as well as severity and progression of the disease.

This study shed light on the impact of non-dopaminergic brain regions in determining disease manifestation and severity, offering critical insights into disease progression. These findings are critical for predicting individual trajectories of Parkinson’s disease and underscore the importance of personalized approaches in managing this complex neurodegenerative disorder.

Future preclinical studies should evaluate the impact of genetic risk factors and comorbidities that may influence the development of these different clinical and biological subgroups in PD.

## Data Availability

All data produced in the present study are available upon reasonable request to the authors.

https://ida.loni.usc.edu/

## Acknowledgment

The authors report no disclosures relevant to the manuscript. SPC is supported by #NEXTGENERATIONEU (NGEU) and funded by the Ministry of University and Research (MUR), National Recovery and Resilience Plan (NRRP), project MNESYS (PE0000006) – A multiscale integrated approach to the study of the nervous system in health and disease (DN. 1553 11.10.2022). We thank Maxine Paige Pritchett for the careful revision of the English language in our manuscript.

## Authors’ roles

R. M., C. M., A. G., M. I., and P. D. conducted the statistical analyses and interpreted the data. S. P. C., R. M., and C. M. drafted the manuscript. L. G., P. M., C. Z., and A. L. collected data and revised the manuscript. A. Pi., A. Pa., M. A., E. M. V., and C. T. contributed to data interpretation. All authors reviewed and approved the final manuscript. C.T. and S. P. C. was responsible for the study’s design and conceptualization.

