## Supplementary materials for "DAT-SPECT profiling for biological definition of two prospective Parkinson’s disease cohorts"

**Methods**

**Study participants**

The study sample comprised 249 de novo Parkinson's disease (PD) patients selected from the Parkinson's Progression Markers Initiative (PPMI) database. All participants met the following inclusion criteria: (1) a clinical diagnosis of idiopathic PD based on the diagnostic criteria of the International Parkinson and Movement Disorder Society; (2) disease duration of ≤ 2 years; (3) absence of prior treatment; and (4) availability of a ^123^I-FP-CIT SPECT scan to assess brain dopaminergic density. Patients with moderate or advanced PD (Hoehn and Yahr stage ≥ III), a neurological diagnosis of mild cognitive impairment (MCI) or dementia at baseline, or a follow-up duration of ≤ 1 year were excluded.

**Clinical and neuropsychological assessment**

All PPMI subjects were administered a comprehensive clinical motor and non-motor assessment.

*Motor clinical features* - The motor evaluation included the MDS‐UPDRS part II, III and IV which respectively assess the motor difficulties in daily life and the global motor burden.^1^ Based on MDS-UPDRS part III, we divided the severity of motor impairment into tremor (sum of items 15–18), rigidity (item 3), bradykinesia (sum of items 2, 4–9 and 14) and postural (sum of items 1 and 9–13).^2^

As the dopaminergic therapy for PD patients is highly heterogeneous, we considered the levodopa equivalent daily dose (LEDD) for every participant. Total LEDD is defined as the cumulative exposure of patients to all dopaminergic drugs. We have also included information on patients who have undergone advanced therapy protocols, specifically deep brain stimulation (DBS).

*Non-motor clinical features* - The non-motor assessment, instead, included the MDS-UPDRS part I, which evaluates the non-motor aspects of daily living experiences, the Scales for Outcomes in PD-Autonomic (SCOPA-AUT),^3^ which assesses autonomic dysfunction, the REM (rapid eye movement) sleep behaviour disorder-screening questionnaire (RBDSQ) that verifies the presence of sleep disturbances,^4^ with RBD disorders defined as a score ≥ 6.^5^ The neuropsychiatric evaluation include the Geriatric Depression Scale (GDS)^6^, Questionnaire for Impulsive-Compulsive Disorders in Parkinson's Disease (QUIP-Current-Short) (QUIPCS)^7^, a scale designed to assess the presence and severity of impulse control disorder (ICD)^8^ and related disorders in PD and both state trait anxiety inventory (STAI) subscales: STAI state (STAI-S) and STAI trait (STAI-T).^9^

*Cognitive profile* - Global cognition was assessed through Montreal Cognitive Assessment (MoCA).^10^ Other cognitive tests considered included the Benton Judgment of Line Orientation (JOLO)^11^ which allows to assess visuo-spatial abilities and the Hopkins Verbal Learning Test (HVLT), a brief word-learning task that evaluates, over the course of multiple trials, verbal learning and memory providing information about both immediate and delayed recall abilities.^12^ Participants were also asked to complete the symbol digit modalities test (SDM),^13^ a test measuring information processing speed, the letter number sequencing (LNS),^14^ to assess working memory abilities and the semantic fluency test.^15^

The longitudinal changes in the MoCA were calculated by determining individual slopes for each participant (MoCA slope).^16^ These slopes were derived using linear regression models adjusted for age, sex, and education.

**CSF biomarker**

Baseline samples for α-syn seeding aggregation activity (SAA) (one sample per participant) were downloaded from the PPMI biospecimen database. Samples were categorized as positive for α-syn SAA if all three replicates were positive, negative if zero or one replicate showed positivity, or inconclusive if two replicates were positive. The Aprion α-syn SAA assay, developed by Concha-Marambio et al.^17^, was previously described following a detailed protocol.^18^

Data on the levels of α-syn ^19^, Aβ_1-42_ (Aβ_1-42_), total Tau (t-Tau), phosphorylated Tau (p-Tau_181_),^20^ and neurofilament light chain (NfL) in the CSF were also analyzed. Ratios between α-syn, Aβ_1-42_, t-Tau, and p-Tau_181_ CSF biomarkers were calculated following the guidelines outlined by Kang et al.^20^

**Sensitivity analysis**

*Participants* - The inclusion and exclusion criteria applied are the same used for the PPMI cohort. The validation dataset included 84 participants (mean age ± SD: 64.55 ± 9.4 years; sex [M/F]: 52/32). Written informed consent was obtained from each participant. The study was conducted in accordance with the Declaration of Helsinki.

*Clinical evaluation* - Participants underwent comprehensive clinical evaluations - including a physical examination and assessments of motor, non-motor and cognitive symptoms - along with a baseline *DAT-SPECT* acquisition*,* followed by a longitudinal follow-up (mean duration: 5.27 ± 2.4 years). Motor symptoms severity was assessed using the MDS-UPDRS part III, while non-motor symptoms were evaluated with the REM (rapid eye movement) sleep behavior disorder-screening questionnaire (RBDSQ) and global cognition was measured using the Mini-Mental State Examination (MMSE).^21^

*DAT-SPECT	acquisition and pre-processing* - Brain *DAT-SPECT* acquisition was performed 3 hours after tracer administration using a Discovery 630 scanner (General Electric, Milwaukee, WI). Images were reconstructed via filtered back-projection, with a three-dimensional Butterworth post-filter (order 10.0; cut-off 0.50 cycles/cm) and corrected for attenuation using Chang's method (attenuation coefficient 0.15 cM^1^).

*Statistical analysis* - We performed proportion tests, Chi-square tests and one-way ANOVA to compare demographic characteristics between the independent cohorts (PPMI cohort vs validation cohort). Subsequently, a two-step cluster analysis was performed on the validation cohort to validate the findings from the PPMI patient group. See **Fig. 1** for a schematic representation. The performed clustering analysis mimics the procedure adopted for the PPMI cohort, thus considering first the *DAT-SPECT* SBR values of left and right caudate and putamen and total MDS-UPDRS III score, as well as the presence of deficit in motor sub-scores.

Chi-square tests and non-parametric Kruskal-Wallis tests were performed to compare characteristics within the validation cohort, with the significance threshold set at 0.05.

A mixed linear model was employed to estimate the rate of longitudinal change in LEDD and MDS-UPDRS part III across the validation subgroups, adjusting for age, sex and education.

**Results**

**Sensitivity analysis**

The two independent cohort (PPMI and validation cohorts) were comparable in terms of numerosity, baseline age, and sex.

At baseline, the D+/D clusters were comparable in terms of age of onset, disease duration and education level, and a greater male prevalence in the D cluster (**Supplementary Table 12)**. The four subgroups of the validation cohort, at baseline, were comparable in terms of age at onset and disease duration; however, the D/M subgroup exhibited higher education levels and a greater presence of males than the other subgroups (**Supplementary Table 13**).

*Clinical features* ***-*** In line with the findings from the PPMI cohort, the D+/M+ and D/M+ subgroups exhibited significantly higher total scores on part III of the MDS-UPDRS compared to the M subgroups. Specifically, the D+/M+ subgroup demonstrated a higher prevalence of bradykinetic symptoms and a lower incidence of tremor, whereas the D/M+ subgroup showed a greater predominance of rigidity-related symptoms (**Supplementary Table 13**).

*Longitudinal assessment* - The estimated annual rate of change in the total MDS-UPDRS Part III score was found to be significantly higher in the D+/M+ subgroup (6.2 points per year), as found in the PPMI cohort. Additionally, a trend not significant was observed in the longitudinal changes in LEDD values of D+/M+ compared to other subgroups (**Supplementary Table 13**).

**Supplementary data**

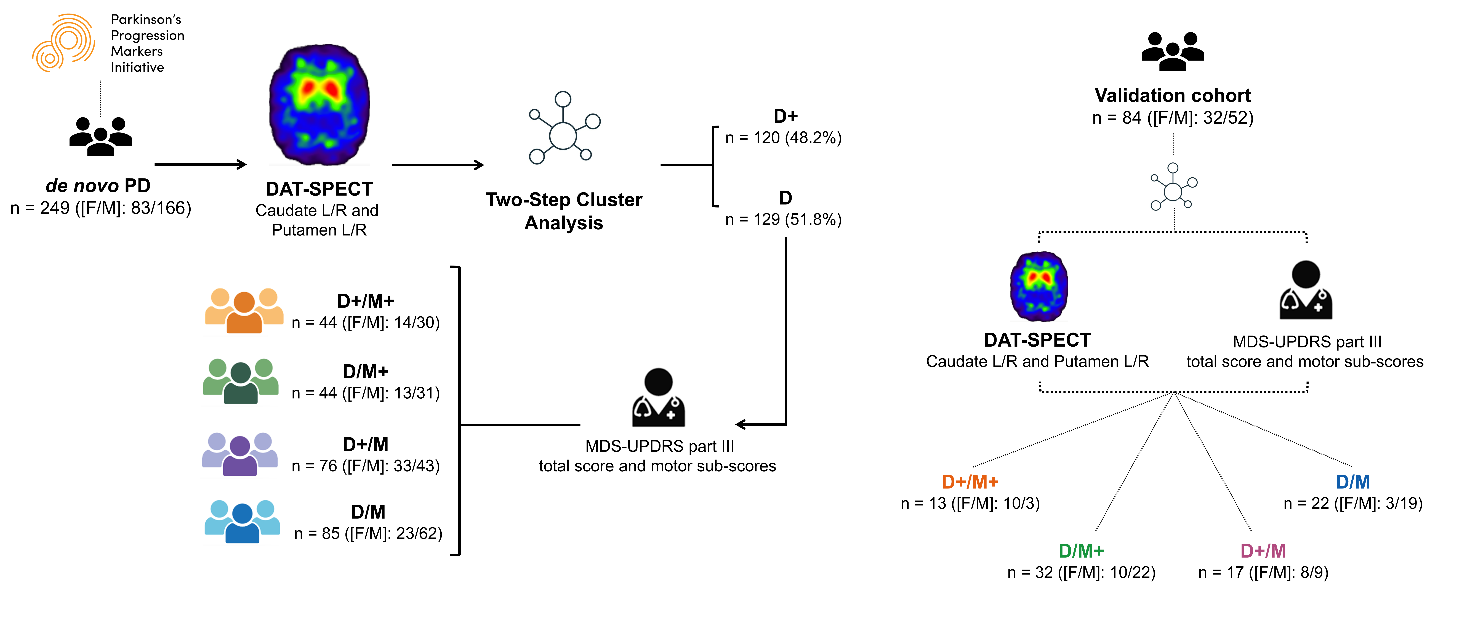

**Supplementary Fig. 1 - Methodological Framework of Cluster Analysis.**

Classification of a de novo Parkinson's disease (PD) cohort (n = 249) using DAT-SPECT imaging and MDS-UPDRS-III motor assessments, followed by Two-Step Cluster Analysis to define distinct subgroups. The same procedure was performed on a validation cohort (n=84).

Abbreviations: Parkinson’s disease (PD); Movement Disorders Society - Unified Parkinson’s Disease Rating Scale (MDS-UPDRS); Dopamine Active Transporter (DAT); Single Photon Emission Computerized Tomography (SPECT); Left (L); Right (R).

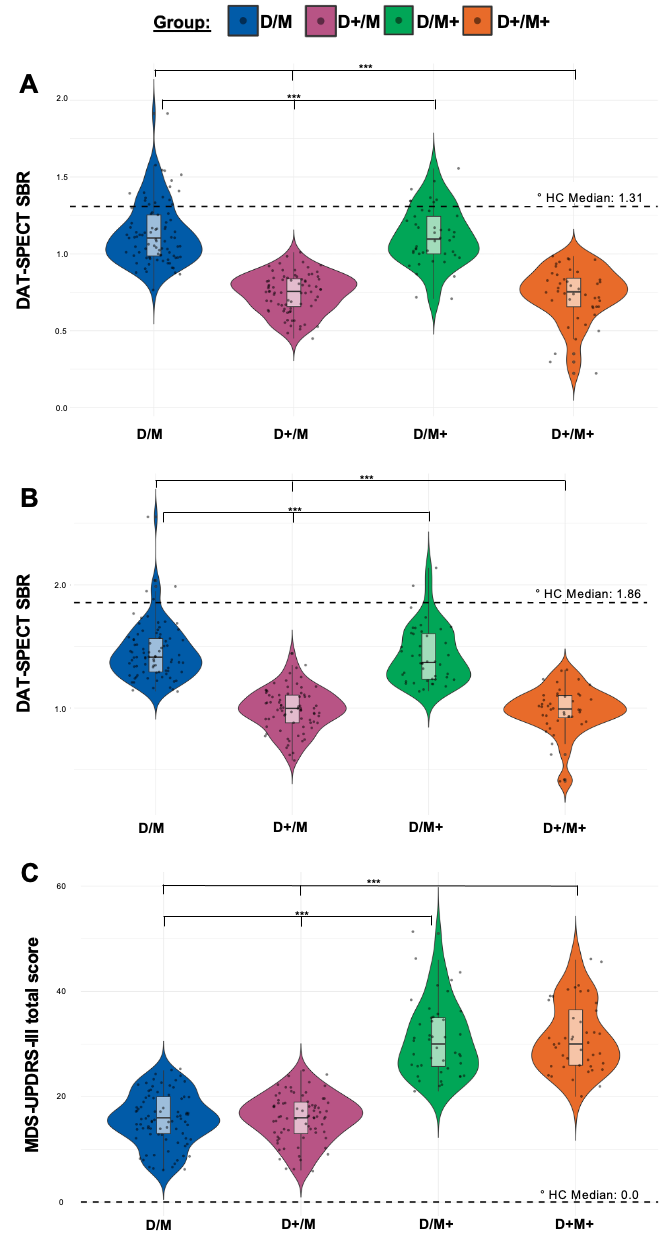

**Supplementary Fig. 2 - Clustering solution of the considered PPMI cohort**

Identification of 4 subgroups by Two-Steps Clustering analysis by (**A**) caudate *DATSCAN* uptake, (**B**) putamen *DATSCAN* uptake and (**C**) MDS-UPDRS III total score.

Abbreviations: Dopamine Active Transporter (DAT); Single Photon Emission Computerized Tomography (SPECT); Specific Binding Ratio (SBR); Healthy Controls (HC); Movement Disorders Society - Unified Parkinson’s Disease Rating Scale (MDS-UPDRS).

Significance: *p ≤ .05; ** p ≤ .01; *** p ≤.001

° Subgroups differ significantly from HC (p < 0.001)

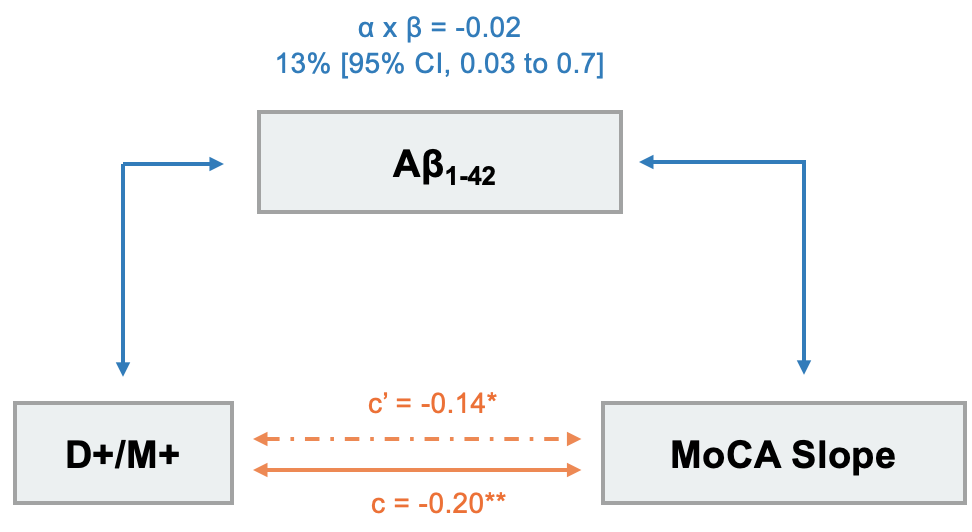

**Supplementary Fig. 3 - Mediation Analysis**

The direct effect (c') measures MoCA slope change per 1-unit D+/M+ increase, controlling for Aβ1-42. The indirect effect (α × β) captures MoCA slope changes via Aβ1-42 alterations. The total effect (c) combines direct and indirect effects.

**Significance:** *p ≤ .05; **p ≤ .01; ***p ≤ .001

**Supplementary Table 1** – A Four-Step Method for Testing Mediation

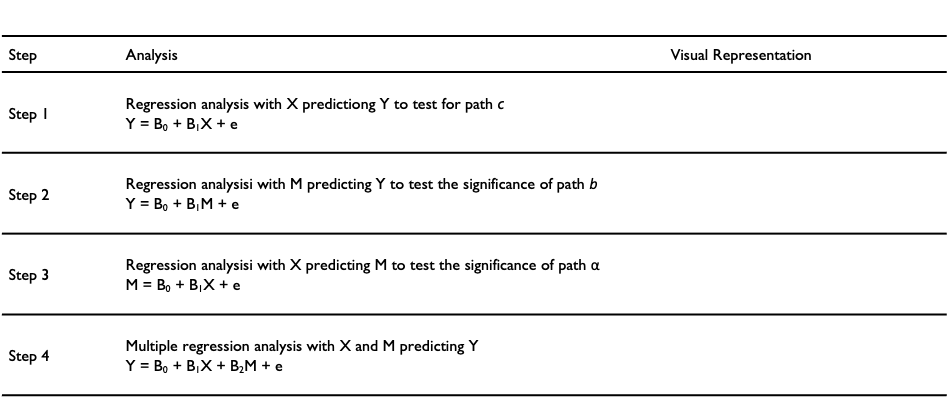

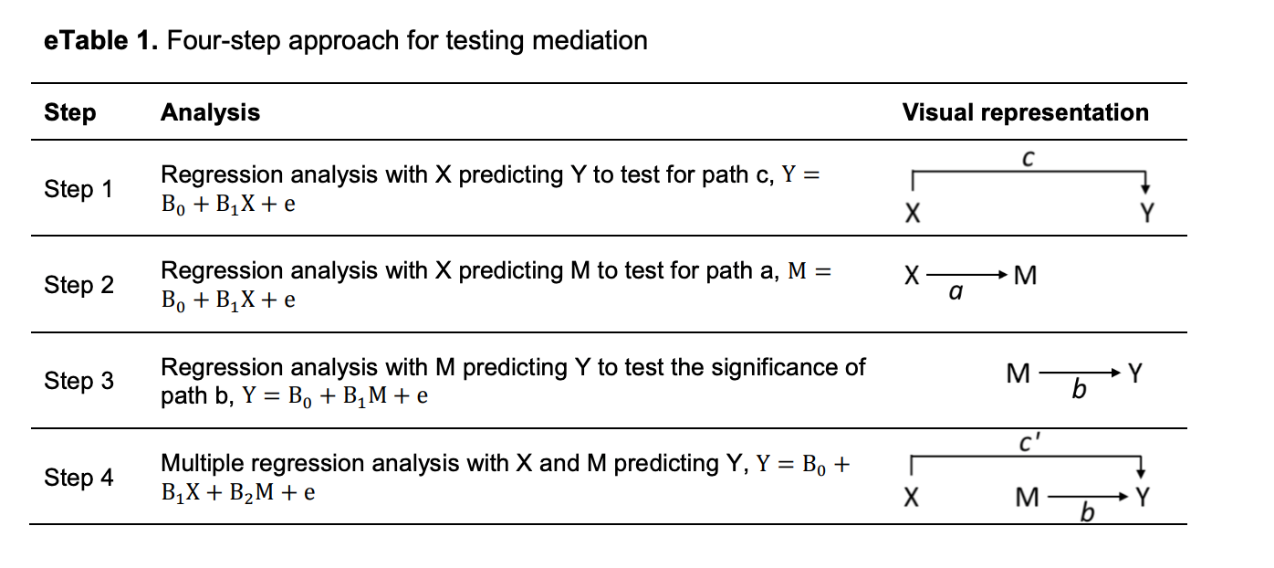

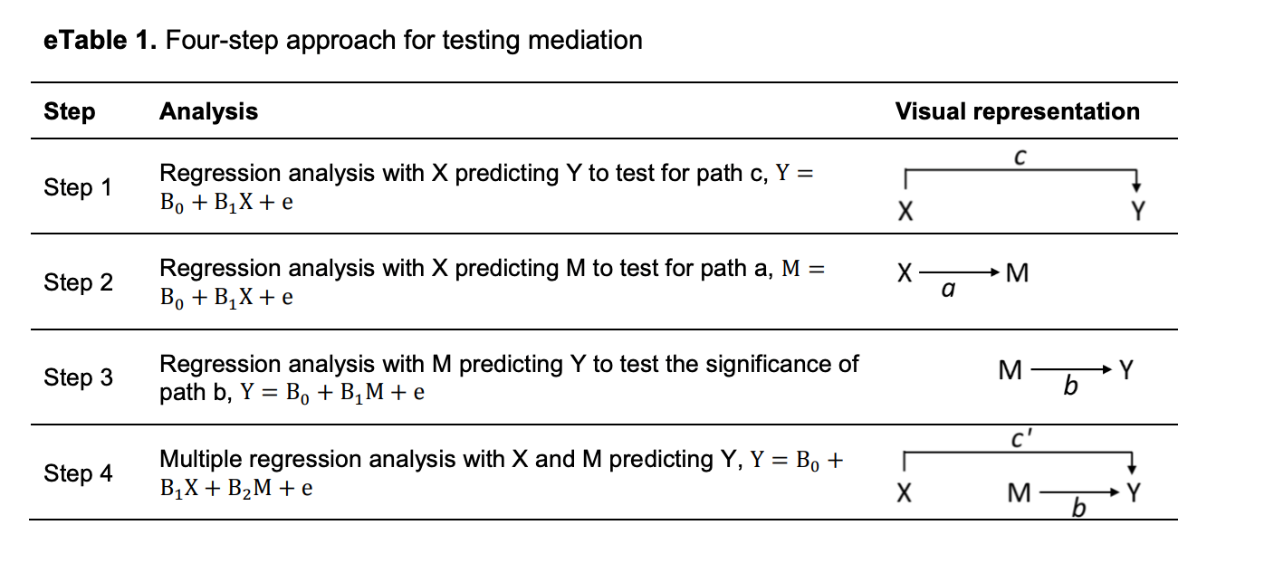

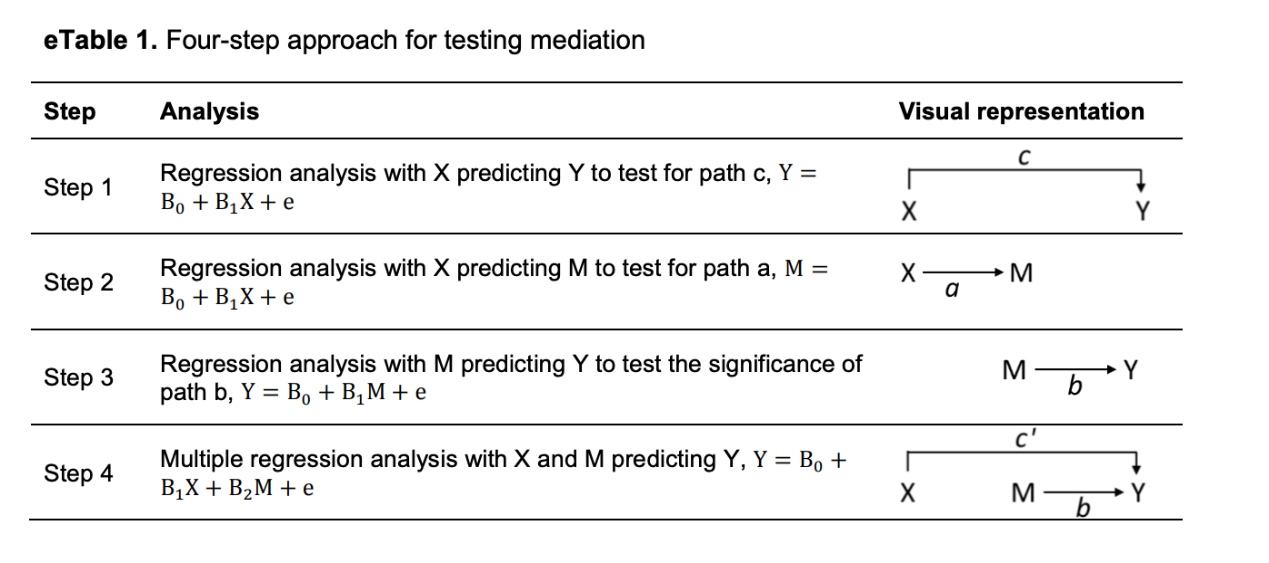

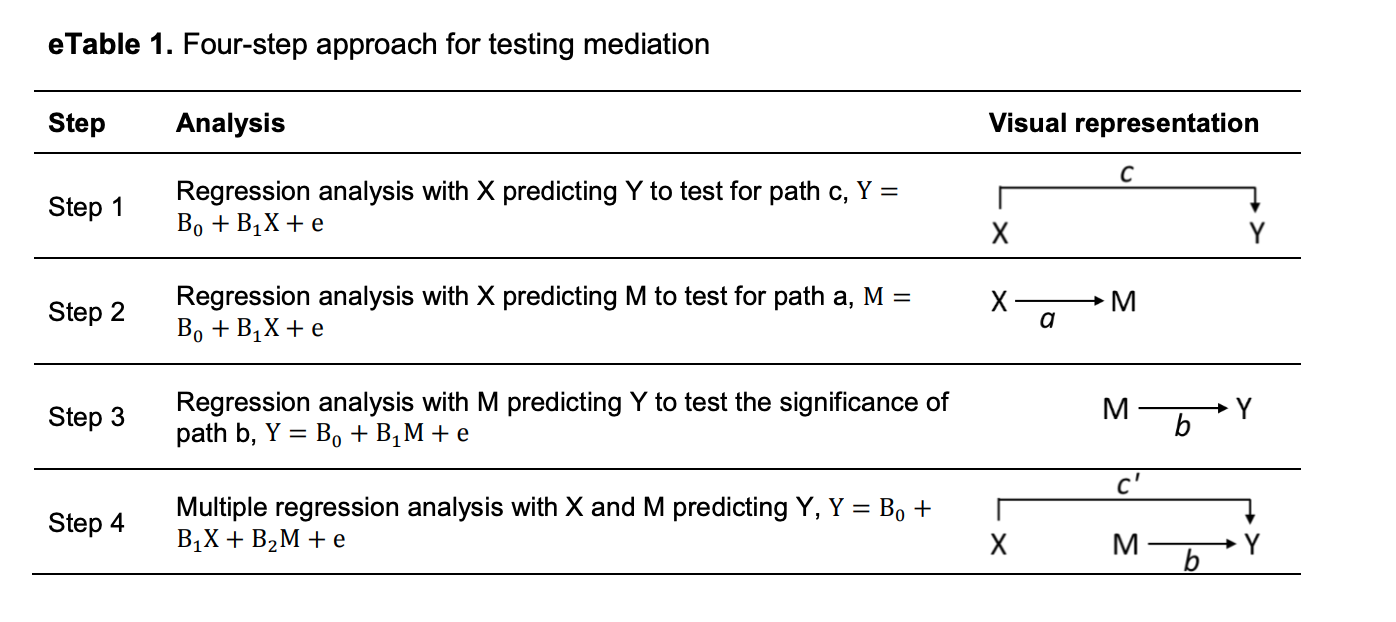

**Supplementary Table 2 -** Summary of clustering analysis

|  | **D+** | **D** | **p-value** |
| --- | --- | --- | --- |
| N (%) | 120 (48.2%) | 129 (51.8%) | - |
| ***Brain imaging*** | | | |
| Cautate L/R | 0.7 (0.1) | 1.1 (0.2) | **< 0.001** § |
| Putamen L/R | 1.0 (0.2) | 1.4 (0.2) | **< 0.001** § |
| Ratio c/p | 0.8 (0.1) | 0.8 (0.1) | 0.078 |
| AI | 5.7 (3.9) | 6.0 (3.4) | 0.309 |
| Delay between first clinical evaluation and SPECT acquisition (months) | 1.2 (1.0) | 1.1 (1.4) | 0.917 |
|  | **M+** | **M** | **p-value** |
| N (%) | 88 (35.3%) | 161 (64.7%) | - |
| ***Clinical assessment*** | | | |
| MDS-UPDRS III | 31.1 (6.8) | 15.82 (4.7) | **< 0.001** § |
| Severity of motor signs (MDS-UPDRS-III) | | |  |
| *Bradykinesia (tot. score range: 0-56)* | 17.1 (4.6) | 7.5 (3.1) | **< 0.001** § |
| *Rigidity (tot. score range: 0-20)* | 6.3 (2.5) | 2.5 (1.6) | **< 0.001** § |
| *Resting tremor (tot. score range: 0-40)* | 5.0 (3.6) | 4.3 (2.9) | **< 0.001** § |
| *Postural (tot. score range: 0-24)* | 3.1 (2.0) | 1.5 (1.2) | 0.165 |

Linear regression models (with Benjamini-Hochberg post-hoc correction) were performed to compare brain imaging and clinical features between clusters (D+/D- and M+/M-). Benjamini-Hochberg correction was applied to control for multiple comparisons, calculated separately for brain imaging, and for clinical assessment.

Abbreviations: Movement Disorders Society - Unified Parkinson’s Disease Rating Scale (MDS-UPDRS).

Significant p-values (p<0.05) are reported in bold.

§ Significance survived Benjamini-Hochberg’s correction.

**Supplementary Table 3** - Comparison of baseline demographic between D+/D clusters

|  | **D+** | **D** | **p-value** |
| --- | --- | --- | --- |
| N (%) | 120 (48.2%) | 129 (51.8%) | - |
| Age at baseline | 62.8 (9.3) | 63.1 (8.8) | 0.756 |
| Age at onset | 61.0 (9.6) | 61.0 (8.8) | 0.995 |
| Age DAT-SPECT | 63.2 (9.3) | 63.5 (8.8) | 0.775 |
| Age MRI | 63.3 (8.7) | 63.2 (8.9) | 0.939 |
| Age at the start of therapy (LEDD) | 68.7 (8.7) | 69.0 (9.0) | 0.823 |
| Sex (% of males) | 73 (60.8%) | 93 (72.1%) | 0.080 |
| Education (years) | 15.5 (2.8) | 15.7 (2.8) | 0.525 |
| Disease Duration (years) | 0.2 (0.4) | 0.2 (0.5) | 0.473 |
| LEDD at baseline | 0.0 (0.0) | 0.0 (0.0) | - |
| DBS | 6 (5%) | 3 (2.3%) | 0.429 |
| Time between first clinical evaluation and DBS (years) | 9.3 (2.7) | 8.6 (3.0) | 0.502 |
| ***Ethnicity*** | | | |
| Hispanic/Latin | 1 (0.8%) | 2 (1.6%) | 1.000 |
| White | 114 (95%) | 126 (97.7%) | 0.429 |
| ***Family history*** | | | |
| Positive family history (%) | 22 (18.3%) | 38 (29.5%) | 0.057 |
| Positive family history-first generation (%) | 11 (9.2%) | 19 (14.7%) | 0.249 |

Comparison of baseline demographics between clusters using Chi-Squared and One-Way ANOVA (significance level of 0.05).

Abbreviations: Deep Brain Stimulation (DBS); Levodopa equivalent daily dose (LEDD); Number (N), Magnetic Resonance Imaging (MRI), Dopamine Active Transporter (DAT); Single Photon Emission Computerized Tomography (SPECT).

Significant p-values (p<0.05) are reported in bold.

**Supplementary Table 4** - Comparison of the baseline clinical features between D+/D clusters

|  | **D+** | **D** | **p-value** |
| --- | --- | --- | --- |
| N (%) | 120 (48.2%) | 129 (51.8%) | - |
| ***Clinical assessment*** | | | |
| MDS-UPDRS II | 5.8 (3.7) | 5.7 (4.0) | 0.668 |
| MDS-UPDRS III | 21.6 (9.1) | 20.9 (9.2) | 0.445 |
| Severity of motor signs (MDS-UPDRS-III) | | |  |
| *Bradykinesia (tot. score range: 0-56)* | 11.4 (6.1) | 10.5 (5.6) | 0.2857 |
| *Rigidity (tot. score range: 0-20)* | 3.8 (2.6) | 3.9 (2.8) | 0.948 |
| *Resting tremor (tot. score range: 0-40)* | 4.4 (2.8) | 4.7 (3.4) | 0.563 |
| *Postural (tot. score range: 0-24)* | 2.3 (1.8) | 1.9 (1.6) | **0.044** |
| Clinical subtypes (*****) | | | 0.369 |
| *Mild Motor-Predominant* | 60 (50%) | 75 (58.1%) | - |
| *Intermediate* | 43 (35.8%) | 36 (27.9%) | - |
| *Diffuse Malignant* | 17 (14.2%) | 18 (14%) | - |
| ***Non-motor symptoms and signs*** | | | |
| MDS-UPDRS I | 1.3 (1.5) | 1.3 (1.9) | 0.746 |
| *Hallucinations and psychosis* | 0.02 (0.1) | 0.04 (0.2) | 0.351 |
| *Apathy* | 0.2 (0.4) | 0.3 (0.6) | **0.020** |
| *Pain* | 0.7 (0.8) | 0.8 (0.9) | 0.351 |
| *Constipation* | 0.5 (0.7) | 0.4 (0.6) | 0.352 |
| *Fatigue* | 0.6 (0.8) | 0.7 (0.8) | 0.432 |
| SCOPA-AUT | 10.9 (6.6) | 10 (5.8) | 0.229 |
| *Orofacial* | 1.0 (1.1) | 1.1 (1.3) | 0.585 |
| *Constipation* | 1.2 (1.4) | 1.0 (1.2) | 0.175 |
| *Urinary* | 4.3 (3.1) | 4.1 (2.7) | 0.419 |
| *Cardiovascular* | 0.5 (0.8) | 0.4 (0.8) | 0.107 |
| *Pupillo-Motor* | 0.3 (0.5) | 0.4 (0.7) | 0.176 |
| *Thermoregulatory* | 1.2 (1.5) | 1.0 (1.2) | 0.308 |
| *Sexual* | 1.3 (1.7) | 1.2 (1.6) | 0.474 |
| RBDSQ | 3.2 (2.9) | 3.3 (2.6) | 0.935 |
| RBD disorders (RBDsq ≥ 6) | 23 (19.2%) | 24 (18.6%) | 0.693 |
| QUIPCS | 3.5 (1.8) | 3.5 (1.7) | 0.949 |
| GDS | 5.2 (1.7) | 5.1 (1.4) | 0.816 |
| STAI S | 45.9 (8.9) | 48 (4.7) | **0.013** |
| STAI T | 45.1 (8.2) | 45.9 (4.0) | 0.216 |
| ***Cognitive assessment*** | | | |
| MoCA (at baseline) | 26.8 (2.8) | 27.0 (2.3) | 0.175 |
| MoCA Slope | -0.4 (0.9) | -0.2 (0.7) | **0.028** |
| BJLO | 13.0 (2.4) | 13.1 (2.7) | 0.988 |
| LNS | 11.4 (2.8) | 11.9 (2.6) | 0.152 |
| Semantic fluency | 49.6 (9.9) | 51.6 (9.6) | 0.097 |
| HVLT, immediate recall | 44.6 (11.0) | 46.6 (10.3) | **0.041** |
| HVLT, delayed recall | 43.8 (11.2) | 45.2 (11.1) | 0.123 |
| HVLT, retention | 47.2 (11.7) | 46.7 (11.5) | 0.998 |
| HVLT, recognition index | 42.4 (11.9) | 46.8 (10.7) | **< 0.001** § |
| SDM | 44.2 (9.0) | 45.9 (9.3) | 0.058 |

Linear and logistic regression (with the Benjamini-Hochberg post hoc correction) were performed to compare clinical features at baseline between groups. Benjamini-Hochberg correction was applied to control for multiple comparisons, calculated separately for clinical motor, non-motor and cognitive symptoms.

Missing value: MDS-UPDRS II (n=1); SCOPA-AUT - RBDsq - STAI - GDS – QUIP-cs (n=3); BJLO - LNS - Semantic fluency - HVLT - SDM (n=4).

Abbreviations: Benton Judgment of Line Orientation (BJLO); Hopkins Verbal Learning Test-Revised (HVLT); Letter-Number Sequencing (LNS); Symbol Digit Modality (SDM); Geriatric Depression Scale (GDS); Montreal Cognitive Assessment (MoCA); Movement Disorders Society - Unified Parkinson’s Disease Rating Scale (MDS-UPDRS); Number (N); Questionnaire for Impulsive-Compulsive Disorders in Parkinson’s Disease (QUIP-cs); REM Sleep Behavior Disorder Screening Questionnaire (RBDsq); Scales for Outcomes in PD Autonomic (SCOPA-AUT); State-Trait Anxiety Inventory-State (STAI-S); State-Trait Anxiety Inventory-Trait (STAI-T).

(*****) Indicates variables analyzed using the chi-square test.

Significant p-values (p<0.05) are reported in bold.

§ Significance survived Benjamini-Hochberg’s correction.

**Supplementary Table 5** - Comparison of CSF biomarkers between D+/D clusters

|  | **D+** | **D** | **p-value** |
| --- | --- | --- | --- |
| N (%) | 120 (48.2%) | 129 (51.8%) | - |
| ***Biomarkers*** | | | |
| SAA (*****) |  |  | 0.894 |
| *Inconclusive* | 2 (1.8%) | 2.0 (1.6%) | - |
| *Positive* | 103 (92%) | 115 (93.5%) | - |
| *Negative* | 7 (6.3%) | 6 (4.9%) | - |
| α-syn | 1437.9 (685.9) | 1631.3 (719.9) | **0.027** |
| Aβ_1-42_ | 865.6 (433.0) | 965.6 (448.4) | 0.066 |
| p-Tau_181_ | 13.9 (4.5) | 15.7 (5.8) | **0.012** |
| t-Tau | 158.4 (52.2) | 180.5 (61.6) | **0.003** § |
| NfL | 13.7 (6.1) | 13.4 (8.2) | 0.710 |
| p-Tau_181_/α-syn | 0.08 (0.01) | 0.09 (0.01) | **0.042** |
| t-Tau/α-syn | 0.1 (0.02) | 0.1 (0.03) | 0.189 |
| Aβ_1-42_/α-syn | 0.6 (0.2) | 0.6 (0.2) | 0.625 |
| p-Tau_181_/Aβ_1-42_ | 0.009 (0.002) | 0.01 (0.002) | 0.093 |
| t-Tau/Aβ_1-42_ | 0.2 (0.1) | 0.2 (0.1) | 0.152 |
| p-Tau_181_/t-Tau | 0.1 (0.01) | 0.1 (0.01) | 0.148 |

Linear and logistic regression (with the Benjamini-Hochberg post hoc correction) were performed to compared CSF biomarkers at baseline between clusters at significance level of 0.05. Benjamini-Hochberg correction was applied to control for multiple comparisons.

Missing value: SAA (n=14); α-syn (n=11); t-Tau (n=17); p-Tau_181_ (n=30); Aβ_1-42_ (n=14); NfL (n=11).

Abbreviations: Amyloid-β_1-42_ (Aβ_1-42_); Asymmetry Index (AI); α-synuclein (α-syn); caudate/putamen ratio (c/p); Cerebrospinal Fluid (CSF); Neurofilament light Chain (NfL); Number (N); Phosphorylated Tau (p-Tau_181_); Single Photon Emission Computerized Tomography (SPECT); total Tau (t-Tau)

(*****) Indicates variables analyzed using the chi-square test.

Significant p-values (p<0.05) are reported in bold.

§ Significance survived Benjamini-Hochberg’s correction.

**Supplementary Table 6 -** Models Comparing Rate of Change in LEDD, MDS-UPDRS III Score and motor domains, MDR-UPDRS IV and motor complications (*Model A*) and cognitive abilities (*Model B*) between D+/D clusters

|  | **D** | |
| --- | --- | --- |
| Reference: **D+** | Estimate (95% CI) | p-value |
| ***Rate of Change in Motor Score*** | |  |
| LEDD | -28.80 (-65.5 to 7.9) | 0.124 |
| MDS-UPDRS-III score | -0.55 (-2.7 to 1.6) | 0.613 |
| *Bradykinesia sub-score* | -1.55 (-2.8 to -0.3) | **0.018** |
| *Tremor sub-score* | 0.83 (0.1 to 1.53) | **0.023** |
| *Rigidity sub-score* | 0.25 -0.3 to 0.8) | 0.390 |
| *Postural sub-score* | -0.52 (-1 to .0.06) | **0.027** |
| THWD sub-score | -0.49 (-1.3 to 0.3) | 0.222 |
| THOFF sub-score | -0.73 (-1.3 to -0.1) | **0.017** |
| ***Rate of Change in Cognitive Score*** | |  |
| *MoCA score* | 0.35 (-0.3 to 1) | 0.285 |
| *BJLO* | 0.53 (-0.03 to 1.1) | 0.063 |
| *HVLT* | 2.39 (0.2 to 4.5) | **0.029** |
| *LNS* | 0.52 (-0.1 to 1.1) | 0.089 |
| *Semantic Fluency* | 1.42 (-0.7 to 3.5) | 0.192 |
| *SDM* | 1.55 (-0.5 to 3.6) | 0.145 |

The longitudinal comparison between clusters was performed using a linear mixed-effects model, with age, sex, and years of education as covariates.
Abbreviations: Benton Judgment of Line Orientation (BJLO); Confidence Interval (CI); Hopkins Verbal Learning Test (HVLT); Letter Number Sequencing (LNS); Movement Disorder Society – Unified Parkinson’s Disease Rating Scale (MDS-UPDRS); Montreal Cognitive Assessment (MoCA); Symbol Digit Modality (SDM); Total Hours with Dyskinesia (THWD); Total Hours OFF (THOFF)

Significant p-values (p<0.05) are reported in bold.

**Supplementary Table 7** - Comparison of baseline demographic and genetic characteristics among the four subgroups of Parkinson’s disease population.

|  | **D+/M+** | **D/M+** | **D+/M** | **D/M** | **p-value** |
| --- | --- | --- | --- | --- | --- |
| N (%) | 44 (17.7%) | 44 (17.7%) | 76 (30.5%) | 85 (34.1%) | - |
| Age at baseline (*****) | 64.4 (9.2) | 64.8 (8.2) | 61.8 (9.3) | 62.3 (9.1) | 0.203 |
| Age at onset | 62.9 (9.7) | 62.1 (8.3) | 59.9 (9.5) | 60.4 (9.0) | 0.368 |
| Age DAT-SPECT | 64.9 (9.3) | 65.1 (8.1) | 62.3 (9.2) | 62.7 (9.1) | 0.211 |
| Age MRI | 65.1 (7.2) | 64.8 (8.5) | 62.3 (9.3) | 62.5 (9.0) | 0.258 |
| Age at the start of therapy (LEDD) | 69.7 (6.4) | 69.7 (7.8) | 68.1 (9.7) | 68.6 (9.6) | 0.706 |
| Sex (% of males) | 30 (68.2%) | 31 (70.5%) | 43 (56.6%) | 62 (72.9%) | 0.150 |
| Education (years) (*****) | 15.87 (2.9) | 15.4 (2.7) | 15.2 (2.7) | 15.8 (2.8) | 0.461 |
| Disease Duration (years) (*****) | 0.18 (0.4) | 0.16 (0.4) | 0.16 (0.4) | 0.24 (0.5) | 0.806 |
| LEDD at baseline | 0.0 (0.0) | 0.0 (0.0) | 0.0 (0.0) | 0.0 (0.0) | - |
| DBS | 6 (13.6 %) | 2 (4.5%) | 11 (14.5%) | 9 (10.6%) | 0.382 |
| Time between first clinical evaluation and DBS (years) | 8.9 (3.0) | 8.7 (1.1) | 9.5 (2.6) | 8.5 (3.3) | 0.384 |
| *Ethnicity* | | | | | |
| Hispanic/Latin | 1 (2.3%) | 0 (0%) | 0 (0%) | 2 (2.4%) | 0.419 |
| White | 43 (97.7%) | 44 (100%) | 71 (93.4%) | 82 (96.5%) | 0.284 |
| *Family history* | | | | |  |
| Positive family history (%) | 10 (22.7%) | 10 (22.7%) | 12 (15.8%) | 28 (32.9%) | 0.086 |
| Positive family history-first generation (%) | 6 (13.6%) | 2 (4.5%) | 5 (6.6%) | 17 (20%) | **0.022** |

Comparison of baseline demographics between subgroups using Chi-Squared and One-Way ANOVA and Kruskal-Wallis H tests (where appropriate) were used (significance level of 0.05).

Abbreviations: Deep Brain Stimulation (DBS); Levodopa equivalent daily dose (LEDD); Number (N), Magnetic Resonance Imaging (MRI), Dopamine Active Transporter (DAT); Single Photon Emission Computerized Tomography (SPECT).

(*****) Indicates variables not normally distributed, for which Kruskal-Wallis non-parametric tests were applied. Significant p-values (p<0.05) are reported in bold.

**Supplementary Table 8** - Comparison of brain imaging and CSF biomarkers among the four subgroups

|  | **D+/M+** | **D/M+** | **D+/M** | **D/M** | **p-value** | | | | | |
| --- | --- | --- | --- | --- | --- | --- | --- | --- | --- | --- |
|  |  |  |  |  | D+/M+ vs D/M+ | D+/M+ vs D+/M | D+/M+ vs D/M | D/M+ vs D+/M | D/M+ vs D/M | D+/M vs D/M |
| N (%) | 44 (17.7%) | 44 (17.7%) | 76 (30.5%) | 85 (34.1%) | - | - | - | - | - | - |
| ***Brain imaging*** | | | | | | | | | | |
| Caudate L/R | 0.7 (0.2) | 1.1 (0.2) | 0.7 (0.1) | 1.1 (0.2) | **< 0.001** § | 0.959 | **< 0.001** § | **< 0.001** § | 0.769 | **< 0.001** § |
| Putamen L/R | 1.0 (0.2) | 1.4 (0.2) | 1.0 (0.2) | 1.5 (0.2) | **< 0.001** § | 0.598 | **< 0.001** § | **< 0.001** § | 0.366 | **< 0.001** § |
| Ratio c/p | 0.8 (0.2) | 0.8 (0.1) | 0.8 (0.1) | 0.8 (0.1) | 0.203 | 0.659 | 0.489 | 0.063 | 0.356 | 0.257 |
| AI | 5.5 (3.8) | 5.7 (3.2) | 5.7 (4.0) | 6.1 (3.6) | 0.742 | 0.782 | 0.659 | 0.553 | 0.988 | 0.327 |
| Delay between first clinical evaluation and SPECT acquisition (months) | 1.06 (1.0) | 0.96 (0.7) | 1.24 (1.0) | 1.22 (1.7) | 0.587 | 0.327 | 0.757 | 0.124 | 0.667 | 0.935 |
| ***Biomarkers*** | | | | | | | | | | |
| SAA (*****) |  |  |  |  |  |  |  |  |  |  |
| Inconclusive | 0 (0.0%) | 2 (4.5%) | 2 (2.6%) | 3 (3.5%) | - | - | - | - | - | - |
| Positive | 36 (81.8%) | 37 (84.1%) | 67 (88.2%) | 78 (92.8%) | - | - | - | - | - | - |
| Negative | 3 (6.8%) | 2 (4.5%) | 4 (5.3%) | 4 (4.7%) | - | - | - | - | - | - |
| α-syn | 1328.3 (463.5) | 1659.1 (737.8) | 1495.6 (774.6) | 1617.3 (714.8) | **0.011** § | 0.178 | **0.021** | 0.328 | 0.983 | 0.377 |
| Aβ1-42 | 762.7 (275.8) | 1047.6 (585.3) | 919.2 (488.6) | 925.1 (359.9) | **0.006** § | 0.073 | **0.018** | 0.235 | 0.165 | 0.975 |
| p-Tau181 | 13.0 (3.4) | 16.5 (6.7) | 14.4 (5.0) | 15.3 (5.4) | **0.005** § | 0.052 | **0.014** | 0.154 | 0.483 | 0.418 |
| t-Tau | 149.7 (46.5) | 190.2 (72.9) | 163.5 (54.9) | 175.8 (55.1) | **0.003** § | 0.110 | **0.006** § | 0.061 | 0.404 | 0.241 |
| NfL | 14.0 (5.2) | 16.4 (11.8) | 13.6 (6.5) | 11.94 (5.3) | 0.240 | 0.316 | 0.193 | 0.362 | **0.027** | **0.035** |
| p-Tau181/α-syn | 0.01 (0.002) | 0.01 (0.002) | 0.01 (0.002) | 0.01 (0.002) | 0.109 | 0.952 | 0.160 | 0.144 | 0.661 | 0.144 |
| t-Tau/α-syn | 0.1 (0.02) | 0.1 (0.02) | 0.1 (0.02) | 0.1 (0.03) | 0.602 | 0.270 | 0.876 | 0.114 | 0.759 | 0.120 |
| Aβ1-42/α-syn | 0.6 (0.1) | 0.7 (0.2) | 0.6 (0.2) | 0.6 (0.2) | 0.105 | 0.370 | 0.732 | 0.381 | 0.170 | 0.643 |
| p-Tau181/Aβ1-42 | 0.02 (0.01) | 0.02 (0.01) | 0.02 (0.005) | 0.02 (0.01) | 0.907 | 0.307 | 0.322 | 0.294 | 0.319 | **0.041** |
| t-Tau/Aβ1-42 | 0.2 (0.1) | 0.2 (0.1) | 0.2 (0.1) | 0.2 (0.1) | 0.713 | 0.136 | 0.528 | 0.317 | 0.242 | **0.030** |
| p-Tau181/t-Tau | 0.1 (0.01) | 0.1 (0.01) | 0.1 (0.01) | 0.1 (0.01) | 0.440 | 0.525 | 0.131 | 0.845 | 0.648 | 0.324 |

Linear and logistic regression models (with the Benjamini-Hochberg post hoc correction) were performed to compare brain imaging and CSF biomarkers at baseline between subgroups. Benjamini-Hochberg correction was applied to control for multiple comparisons, calculated for brain imaging and CSF biomarkers.

Missing value: SAA (n=14); α-syn (n=11); t-Tau (n=17); p-Tau_181_ (n=30); Aβ_1-42_ (n=14); NfL (n=11).

Abbreviations: AmyloiDβ_1-42_ (Aβ_1-42_); Phosphorylated Tau (p-Tau_181_); total Tau (t-Tau); α-synuclein (α-syn); Neurofilament light Chain (NfL); Number (N); Single Photon Emission Computerized Tomography (SPECT); Asymmetry Index (AI); caudate/putamen ratio (c/p).

(*****) Indicates variables analyzed using the chi-square test. Significant p-values (p<0.05) are reported in bold. § Significance survived Benjamini-Hochberg’s correction.

**Supplementary Table 9** - Models Comparing Rate of Change in LEDD, MDS-UPDRS III Score and motor domains (*Model A*) and MoCA Score (*Model B*) among subgroups of PD patients

|  | **D+/M+** | | **D/M+** | | **D+/M** | | | **D/M** | | |
| --- | --- | --- | --- | --- | --- | --- | --- | --- | --- | --- |
| Reference: **Group** | Estimate (95% CI) | p-value | Estimate (95% CI) | p-value | | Estimate (95% CI) | p-value | | Estimate (95% CI) | p-value |
| ***Rate of Change in Motor Score*** | | | | | | | | | | |
| LEDD | -62.57 (-110.9 to -14.3) | **0.011** | 12.21 (-40.3 to 64.7) | 0.647 | | 8.42 (-30.6 to 47.4) | 0.671 | | 23.88 (-14.5 to 62.3) | 0.222 |
| MDS-UPDRS-III score | -8.60 (-11.2 to -5.9) | **< 0.001** | -8.21 (-10.8 to -5.6) | **< 0.001** | | 4.66 (2.5 to 6.9) | **< 0.001** | | 5.82 (3.7 to 7.9) | **< 0.001** |
| *Bradykinesia sub-score* | -6.23 (-7.7 to -4.7) | **< 0.001** | -4.57 (-6.2 to -2.9) | **< 0.001** | | 2.37 (1.0 to 3.7) | **0.001** | | 4.43 (3.2 to 5.6) | **< 0.001** |
| *Tremor sub-score* | 0.26 (-0.7 to 1.2) | 0.531 | -0.65 (-1.6 to 0.3) | 0.191 | | 0.78 (-0.02 to 1.5) | **0.043** | | -0.54 (-1.3 to 0.2) | 0.152 |
| *Rigidity sub-score* | -1.44 (-2.2 to -0.6) | **< 0.001** | -2.55 (-3.3 to -1.8) | **< 0.001** | | 1.15 (0.5 to 1.8) | **< 0.001** | | 1.33 (0.7 to 1.9) | **< 0.001** |
| *Postural sub-score* | -1.33 (-1.9 to -0.7) | **< 0.001** | -0.46 (-1.1 to 0.1) | 0.141 | | 0.27 (-0.2 to 0.8) | 0.286 | | 0.86 (0.4 to 1.3) | **< 0.001** |
| THWD sub-score | -1.12 (-2.2 to -0.05) | **0.04** | 1.27 (0.3 to 2.3) | **0.013** | | 0.11 (-0.7 to 0.9) | 0.792 | | -0.32 (-1.2 to 0.5) | 0.464 |
| THOFF sub-score | -0.92 (-1.7 to -0.1) | **0.03** | 1.07 (0.2 to 1.9) | **0.011** | | -0.15 (-0.7 to 0.4) | 0.609 | | 0.20 (-0.5 to 0.9) | 0.558 |
| ***Rate of Change in Cognitive Score*** | | | | | | | | | | |
| MoCA score | 1.02 (0.2 to 1.8) | **0.017** | 0.52 (-0.4 to 1.4) | 0.246 | | -0.31 (-1.0 to 0.4) | 0.36 | | -0.69 (-1.4 to -0.1) | **0.047** |
| BJLO | 1.24 (0.50 to 1.98) | **0.001** | 0.67 (-0.1 to 1.4) | 0.084 | | -0.17 (-0.8 to 0.4) | 0.568 | | -0.95 (-1.5 to -0.4) | **0.001** |
| HVLT | 3.17 (0.3 to 6.0) | **0.029** | -1.36 (-4.3 to 1.5) | 0.354 | | 0.65 (-1.7 to 3.0) | 0.582 | | -1.74 (-4.0 to 0.5) | 0.128 |
| LNS | 0.73 (-0.06 to 1.5) | 0.069 | -0.04 (-0.8 to 0.8) | 0.921 | | 0.09 (-0.5 to 0.7) | 0.789 | | -0.55 (-1.2 to 0.1) | 0.085 |
| Semantic Fluency | 1.30 (-1.5 4.1) | 0.371 | -1.44 (-4.3 to 1.4) | 0.321 | | 0.47 (-1.8 to 2.7) | 0.679 | | -0.37 (-2.6 to 1.8) | 0.74 |
| SDM | 2.37 (-0.4 to 5.1) | 0.092 | 0.65 (-2.2 to 3.5) | 0.651 | | 0.18 (-2.1 to 2.4) | 0.871 | | -2.05 (-4.2 to 0.1) | 0.065 |

The longitudinal comparison between subgroups was performed using a linear mixed-effects model, with age, sex, and years of education as covariates.
Abbreviations: Benton Judgment of Line Orientation (BJLO); Confidence Interval (CI); Hopkins Verbal Learning Test (HVLT); Letter Number Sequencing (LNS); Movement Disorder Society – Unified Parkinson’s Disease Rating Scale (MDS-UPDRS); Montreal Cognitive Assessment (MoCA); Symbol Digit Modality (SDM); Total Hours with Dyskinesia (THWD); Total Hours OFF (THOFF)

Significant p-values (p<0.05) are reported in bold.

**Supplementary Table 10 -** Models comparing the rate of change in MDS-UPDRS III score and motor domains conducted pair-to-pair comparisons between subgroups.

|  | **D/M+** | | **D+/M** | | **D/M** | |
| --- | --- | --- | --- | --- | --- | --- |
| Reference group: **D+/M+** | Estimate (95% CI) | p-value | Estimate (95% CI) | p-value | Estimate (95% CI) | p-value |
| ***Rate of Change in Motor Score*** | | | | | | |
| MDS-UPDRS-III score | -0.11 (-2.97 to 2..76) | 0.941 | -10.3 (-12.9 to -7.84) | **< 0.001** | -10.9 (-13.4 to -8.46) | **< 0.001** |
| *Bradykinesia sub-score* | -1.42 (-3.13 to 0.28) | 0.103 | -6.77 (-8.26 to -5.27) | **< 0.001** | -8.03 (-9.49 to -6.57) | **< 0.001** |
| *Tremor sub-score* | 0.75 (-0.51 to 2.01) | 0.242 | -0.26 (-1.37 to 0.84) | 0.641 | 0.59 (-0.49 to 1.67) | 0.283 |
| *Rigidity sub-score* | 0.83 (-0.78 to 1.75) | 0.073 | -2.25 (-3.04 to -1.45) | **< 0.001** | -2.38 (-3.15 to -1.60) | **< 0.001** |
| *Postural sub-score* | -0.71 (-1.47 to 0.49) | 0.067 | -1.30 (-1.96 to 0.49) | **< 0.001** | -1.69 (-2.34 to -1.04) | **< 0.001** |
|  | **D+/M+** | | **D+/M** | | **D/M** | |
| Reference group: **D/M+** | Estimate (95% CI) | p-value | Estimate (95% CI) | p-value | Estimate (95% CI) | p-value |
| ***Rate of Change in Motor Score*** | | | | | | |
| MDS-UPDRS-III score | 0.12 (-2.76 to 2.97) | 0.941 | -10.2 (-12.8 to -7.71) | **< 0.001** | -10.8 (-13.3 to -8.33) | **< 0.001** |
| *Bradykinesia sub-score* | 1.42 (-0.29 to 3.13) | 0.103 | -5.35 (-6.86 to -3.84) | **< 0.001** | -6.61 (-8.09 to -5.13) | **< 0.001** |
| *Tremor sub-score* | -0.75 (-2.01 to 0.51) | 0.242 | -1.01 (-2.13 to 0.10) | 0.075 | -0.16 (-1.25 to 0.93) | 0.773 |
| *Rigidity sub-score* | -0.83 (-1.75 to 0.08) | 0.073 | -3.08 (-3.88 to -2.28) | **< 0.001** | -3.21 (-3.99 to -2.42) | **< 0.001** |
| *Postural sub-score* | 0.71 (-0.05 to 1.47) | 0.050 | -0.59 (-1.25 to 0.07) | 0.081 | -0.98 (-1.63 to -0.33) | **0.003** |

The comparison of the rate of change in motor domains between subgroups was performed using pair-to-pair comparisons within a linear mixed-effects model, with age, sex, and years of education as covariates.

Abbreviations: Confidence Interval (CI); Movement Disorder Society - Unified Parkinson’s Disease Rating Scale (MDS-UPDRS).

Significant p-values (p<0.05) are reported in bold.

**Supplementary Table 11** – Exploring the Mediation Pathway: The Influence of M on Y (Step 2 Analysis)

|  | **β (95%CI)^a^** | **p-value** |
| --- | --- | --- |
| ***Biomarkers*** | | |
| α-syn | 0.05 (0 to 0) | 0.411 |
| Aβ_1-42_ | 0.15 (0 to 0.1) | **0.019** |
| t-Tau | 0.02 (-0.002 to 0.002) | 0.792 |
| p-Tau_181_ | -0.34 (-0.03 to 0.01) | 0.614 |
| NfL | -0.06 (-0.02 to 0.006) | 0.323 |
| p-Tau_181_/α-syn | -0.18 (-124 to -19) | **0.007** |
| t-Tau/α-syn | -0.14 (-8 to -0.2); | **0.042** |
| Aβ_1-42_ /α-syn | 0.15 (0.8 to 1.2) | **0.025** |
| t-Tau/Aβ_1-42_ | -0.24 (-3.5 to -1) | **< 0.001** |
| p-Tau_181_/ Aβ_1-42_ | -0.25 (-37 to -11) | **< 0.001** |
| p-Tau_181_/t-Tau | -0.09 (-25 to 4) | 0.174 |

Abbreviations: Amyloid-β_1-42_ (Aβ_1-42_); Confidence Interval (CI); Phosphorylated Tau (p-Tau_181_); total Tau (t-Tau); α-synuclein (α-syn); Neurofilament light Chain (NfL); Number (N)

^a^ = β standardised.

Significant p-values (p<0.05) are reported in bold.

**Supplementary Table 12** - Comparison of the baseline clinical features and longitudinal rate of change in LEDD, MDS-UPDRS III Score between D+/D clusters

|  | **D+** | **D** | **p-value** |  |
| --- | --- | --- | --- | --- |
| N (%) | 30 (35.7%) | 54 (64.3%) | - |  |
| Age at baseline | 67.1 (8.2) | 63.1 (9.8) | 0.060 |  |
| Age at onset | 66.3 (8.3) | 62.4 (9.9) | 0.072 |  |
| Sex (% of males) | 11 (36.7%) | 41 (75.9 %) | **0.001** |  |
| Education (years) | 8.0 (4.2) | 9.8 (3.8) | 0.080 |  |
| Disease Duration (years) | 1.4 (0.5) | 1.3 (0.5) | 0.404 |  |
| LEDD at baseline | 0.0 (0.0) | 0.0 (0.0) | - |  |
| ***Clinical assessment at baseline*** | | | |  |
| MDS-UPDRS-III | 14.7 (12.5) | 11.4(7.1) | 0.119 |  |
| *Bradykinesia* | 11 (47.8%) | 20(42.6%) | 0.872 |  |
| *Rigidity* | 16 (69.6%) | 34(72.3%) | 1.000 |  |
| *Resting tremor* | 6 (26.1%) | 19(40.4%) | 0.362 |  |
| *RBD* | 5 (45.5%) | 8 (50.0%) | 1.000 |  |
| MMSE | 27.4 (2.5) | 28.1 (1.5) | 0.226 |  |
| ***Clinical trajectories at follow-up*** | | | | |
| **Rate of Change in Motor Score** | | | | |
|  | LEDD | | MDS-UPDRS-III score | |
| Reference: D+ | Estimate (95% CI) | p-value | Estimate (95% CI) | p-value |
| D | -13.60 (-70.2 to 43.0) | 0.137 | -2.50 (-6.3 to -1.3) | 0.196 |

Chi-square tests and one-way ANOVA were used to compare baseline demographic and clinical data between clusters within the validation cohort. Longitudinal comparisons between groups within the validation cohort were performed using a linear mixed-effects model.

Abbreviations: Levodopa equivalent daily dose (LEDD); Mini-Mental State Examination (MMSE); Movement Disorders Society - Unified Parkinson’s Disease Rating Scale (MDS-UPDRS); Number (N); REM sleep Behaviour Disorder (RBD).

Significant p-values (p<0.05) are reported in bold.

**Supplementary Table 13** - Comparison of the baseline clinical features and longitudinal rate of change in LEDD, MDS-UPDRS III Score among the four subgroups of Parkinson’s disease population

|  | **D+/M+** | **D/M+** | **D+/M** | **D/M** | **p-value** |
| --- | --- | --- | --- | --- | --- |
| N (%) | 13 (15.5%) | 32 (38.1%) | 17 (20.2%) | 22 (26.2%) | - |
| Age at baseline | 68.7 (6.2) | 62.8 (10.6) | 65.9 (9.3) | 63.6 (8.8) | 0.373 |
| Age at onset | 68.2 (6.2) | 62.1 (10.8) | 64.7 (9.4) | 62.8 (8.5) | 0.373 |
| Sex (% of males) | 3 (23.1%) | 22 (68.8%) | 8 (47.1%) | 19 (86.4%) | **0.001** |
| Education (years) | 6.3 (3.9) | 9.6 (4.0) | 9.7 (3.8) | 10.0 (3.5) | **0.014** |
| Disease Duration (years) | 1.2 (0.4) | 1.2 (0.4) | 1.5 (0.5) | 1.3 (0.5) | 0.400 |
| LEDD at baseline | 0 (0) | 0 (0) | 0 (0) | 0 (0) | - |
| ***Clinical assessment at baseline*** | | | | | |
| *MDS-UPDRS-III* | 22.0 (15.7) | 11.7 (6.7) | 9.2 (4.5) | 10.9 (7.8) | **0.022** |
| *Bradykinesia* | 11/11 (100%) | 20/27 (74.1%) | 0/12 (0%) | 0/20 (0%) | **0.000** |
| *Rigidity* | 6/11 (54.5%) | 19/27 (70.4%) | 0/12 (0%) | 0/20 (0%) | **0.000** |
| *Resting tremor* | 4/11 (36.4%) | 19/27 (70.4%) | 12/12 (100%) | 15/20 (75%) | **0.009** |
| *RBD* | 1/6 (16.7%) | 4/8 (50%) | 4/5 (80%) | 4/8 (50%) | 0.218 |
| MMSE | 28.1 (1.9) | 27.7 (1.4) | 26.5 (2.9) | 28.5 (1.5) | 0.172 |
| ***Clinical trajectories at follow-up*** | | | | | |
| **Rate of Change in Motor Score** | | | | | |
|  |  | LEDD | | MDS-UPDRS-III score | |
| Reference: Group |  | Estimate (95% CI) | p-value | Estimate (95% CI) | p-value |
|  | D/M | 10.05 (-50.7 to 70.8) | 0.745 | 20.42 (-1.5 to 6.3) | 0.222 |
|  | D/M+ | 27.85 (-39.8 to 95.5) | 0.418 | 1.57 (-3.1 to 6.2) | 0.501 |
|  | D+/M | 5.11 (-52.8 to 63.0) | 0.862 | 0.21 (-3.5 to 3.9) | 0.910 |
|  | D+/M+ | -55.03 (-127.7 to 17.6) | 0.137 | -6.17 (-11.1 to -1.3) | **0.014** |

Chi-square tests and the non-parametric Kruskal-Wallis test were used to compare baseline demographic and clinical data between subgroups within the validation cohort. The longitudinal comparison between subgroups was performed using a linear mixed-effects model.

Abbreviations: Levodopa equivalent daily dose (LEDD); Mini-Mental State Examination (MMSE); Movement Disorders Society - Unified Parkinson’s Disease Rating Scale (MDS-UPDRS); Number (N); REM sleep Behaviour Disorder (RBD)

Significant p-values (p<0.05) are reported in bold.
